# Smart Discharges improves post-discharge mortality among children with suspected sepsis in Uganda: A prospective before-after study

**DOI:** 10.1101/2025.08.25.25334336

**Authors:** Matthew O Wiens, Cherri Zhang, Vuong Nguyen, Jeffrey N Bone, Elias Kumbakumba, Stephen Businge, Abner Tagoola, Emmanuel Tenywa, Sheila Oyella Sherine, Emmanuel Byaruhanga, Edward Ssemwanga, Jesca Nsungwa, Charles Olaro, J Mark Ansermino, Niranjan Kissoon, Joel Singer, Charles P Larson, Pascal M Lavoie, Dustin Dunsmuir, Peter P Moschovis, Stefanie Novakowski, Jessica Trawin, Clare Komugisha, Bernard T Opar, Mellon Tayebwa, Douglas Mwesigwa, Nicholas West, Nathan Kenya Mugisha, Jerome Kabakyenga

**Affiliations:** Institute for Global Health at BC Children’s and Women’s Hospital, Vancouver, Canada; BC Children’s Hospital Research Institute, Vancouver, Canada; Mbarara Regional Referral Hospital, Mbarara, Uganda; Holy Innocents Children’s Hospital, Mbarara, Uganda; Jinja Regional Referral Hospital, Jinja City, Uganda; Masaka Regional Referral Hospital, Masaka, Uganda; Kawempe National Referral Hospital, Kampala, Uganda; Villa Maria Hospital, Masaka, Uganda; Ministry of Health for the Republic of Uganda, Kampala, Uganda; University of British Columbia, Vancouver, Canada; McGill University, Montréal, Canada; Massachusetts General Hospital, Boston, MA, USA; Walimu, Kampala, Uganda; Mbarara University of Science and Technology, Mbarara, Uganda

**Author notes:** Corresponding authors: Matthew O. Wiens. Institute for Global Health, 305 - 4088 Cambie Street, Vancouver, BC, V5Z 2X8, Canada., Jerome Kabakyenga. Mbarara University of Science and Technology, Mbarara, Uganda.

## Abstract

**Introduction:** Post-discharge mortality among children following an acute illness in low-resource settings is high and demands urgent attention. We aimed to assess the impact of Smart Discharges, a mortality prevention risk-differentiated approach to peri-discharge care among children under five years admitted with suspected sepsis.

**Methods:** We conducted a before-after two-phase study with staggered implementation at six hospitals in Uganda. During a baseline period (Phase-1), clinical prediction algorithms for post-discharge mortality were developed based on clinical and socio-demographic data collected at hospital admission. Outcomes, primarily mortality within six months of discharge, were compared against an interventional period (Phase-2). In the Smart Discharges intervention each family was given soap, a mosquito net, their risk category (low/medium/high/very high), counselling, and educational materials regardless of risk stratification, after which the intensity of recommended follow-up care was determined by age (0-6, 6-60 months) and predicted post-discharge mortality risk.

**Results:** Overall, 13,051 children were enrolled: Phase-1, n=6,955; Phase-2, n=6,096. Characteristics were similar between groups, including mean predicted post-discharge mortality risk (6.3% Phase-1 vs. 5.9% Phase-2). With Smart Discharges, 2,331/3,891 (59.9%) medium/high/very high-risk families attending all their scheduled visits. The observed post-discharge mortality rate was 439 (6.3%) in Phase-1 vs. 296 (4.9%) in Phase-2; adjusted hazard ratio 0.77 (95%CI 0.67 to 0.90) favouring the intervention. In Phase-1, 1,313 (18.9%) children were re-admitted to hospital vs. 1,024 (16.8%) in Phase-2.

**Conclusion:** A simple approach of risk assessment paired with education and engagement through scheduled follow-up after discharge is an appropriate strategy to improve child survival in low-resource settings.

## Introduction

Substantial reductions in childhood mortality have been achieved in recent decades (*1*). Attention has increasingly shifted towards mortality occurring during the post-discharge period (*2–6*). In low-income countries, as many children die within 6 months following hospital discharge as during the initial admission period (*7*). Many of these deaths are likely due to sub-optimal discharge planning, with over half of the deaths occurring at home and approximately half occurring more than one month after hospital discharge (*7*). Thus, plans to improve post-discharge care are urgently required to decrease mortality (*2*).

Few interventional studies, apart from pharmaceutical treatment trials, have targeted post-discharge mortality in low-income country settings. Randomized trials evaluating the effects of micronutrients and iron with antimicrobial prophylaxis have shown no survival benefit during the post-discharge period (*8*). Post-discharge anti-malarial prophylaxis among children with anemia has demonstrated improved survival during the period of prophylaxis (*9*). However, none of these interventions address important socioeconomic and system factors, which are increasingly recognized as major contributors to post-discharge vulnerability (*4,7,10,11*).

Almost half of post-discharge deaths in low-income countries occur at home or in transit to seeking care, suggesting barriers to health-seeking within the formal health system (*7,10*) or a desire to seek care outside of the health system, where effective care of serious illness is unlikely (*12*). Addressing vulnerability through a more informative discharge process and follow-up during the early post-discharge period aims to ensure timely treatment of recurrent illness and improved survival. A previous feasibility study in families of 6-60-month-old children admitted to a Ugandan hospital with a suspected or proven infectious disease indicated that a bundle of interventions, including scheduled follow-up referrals, simple prevention kits, and counseling improved post-discharge health-seeking behaviour and reduced the odds of post-discharge mortality, though the result was not statistically significant (*13*).

Health systems across resource-poor settings lack resilience due to a high disease burden coupled with limited availability of health workers and supplies (*14*). In this setting, precision public health approaches that improve efficiency by targeting the most vulnerable are likely to decrease mortality. Risk-differentiated care specifically targeting those at highest risk may provide a viable and scalable method to improve mortality among children following hospital discharge because post-discharge risk can be reasonably captured through a few key clinical and social features, such as anthropometry, illness severity, access to clean water, and age (*15*).

The objective of this study was to evaluate a risk-differentiated peri-discharge care program in children under five years of age admitted with suspected sepsis in Uganda. We hypothesized that the Smart Discharges program, in which the intensity of follow-up care was determined by age and a risk-stratification algorithm based on simple clinical and socio-demographic data collected at hospital admission, would reduce all-cause six-month post-discharge mortality.

## Methods

### Study Design and Approvals

We conducted a prospective before-after study with staggered implementation to evaluate the impact of a risk-differentiated approach to peri-discharge care on post-discharge mortality among children under 60 months of age who were admitted with suspected sepsis in Uganda. This study was approved by the University of British Columbia (H16-02679, 09-May-2017), the Mbarara University of Science and Technology (No. 15/10-16, 27-Jan-2017), and the Uganda Council of Science and Technology (HS 2207). The study was registered at clinicaltrials.gov (NCT05730452). Written informed consent was obtained from the parent/guardian of all enrolled children. This manuscript adheres to the Transparent Reporting of Evaluations with Nonrandomized Designs (TREND) statement (*16*).

### Study phases: baseline and intervention

The study comprised two phases: Phase-1 was a baseline (control) phase and was also used for developing clinical prediction algorithms of post-discharge mortality (*15*), which were implemented during an interventional period (Phase-2). The epidemiological characteristics of Phase-1 have been published (*7*). At all six sites, baseline Phase-1 was followed by intervention Phase-2, for two age cohorts (0-6 months and 6-60 months) enrolled in parallel. There was no blinding of the intervention among participants’ families, or among the clinicians administering the intervention or assessing the outcomes.

Due to the timing associated with multiple funding sources, recruitment windows differed for the two age cohorts: children aged 0-6 months were enrolled between 15-Jan-2018 and 30-Jun-2023 and children 6-60 months were enrolled from 13-Jul-2017 to 18-Mar-2021 (**Supplementary-S4.1**). COVID-19 associated interruptions prevented a transition of the 0-6 months age group into the intervention Phase-2; thus, we enrolled additional children into Phase 1 between 31-Mar-2020 and 05-Aug-2021. Data were combined across sites and age cohorts for analysis purposes.

### Participants and setting

We enrolled children aged 0-60 months who were admitted from the community with a proven or suspected infection. We have previously demonstrated that about 90% of such children meet the 2005 International Pediatric Consensus Conference (IPSCC) definition, which defines sepsis as the systemic inflammatory response syndrome in the presence of a confirmed or suspected infection (*17,18*). Children were excluded if they resided outside the hospital catchment or were admitted due to trauma or only for short-term observation (<24 hours). Newborns admitted directly after birth (i.e., without having been discharged home) were also excluded.

The study enrolled participants from six hospitals in Uganda: Mbarara Regional Referral Hospital (Southwestern Uganda), Holy Innocents Children’s Hospital (Southwestern Uganda), Masaka Regional Referral Hospital (Central Uganda), and Jinja Regional Referral Hospital (Eastern Uganda); children aged 0-6 months were also enrolled at Villa Maria Hospital (Central Uganda) and Uganda Martyrs Hospital, Ibanda (Southwestern Uganda). In total, these public and faith-based facilities have catchments covering 30 districts with a total population of over 8 million individuals, including about 1.4 million children under 5 years of age during the study period (*19*). These facilities capture both rural and urban populations and provide a reasonable representation of the Ugandan pediatric population outside the capital city of Kampala. The caregivers of all children in both study phases received a bar of soap and a new mosquito net as compensation.

### Outcomes and data collection procedures

The primary outcome was mortality within six months of hospital discharge. Secondary outcomes included number of days to death, hospital readmission, and adherence to the intervention.

Data collection procedures have been described and were identical during both study phases (*7,20*). Briefly, trained study nurses systematically collected data on clinical, social, and demographic variables at admission. Clinical data included anthropometry (to determine nutritional status), vital signs, simple laboratory parameters (glucose, malaria rapid diagnostic test [RDT], HIV RDT, hematocrit, lactate), clinical signs and symptoms, co-morbidities, and healthcare history, including previous hospital admissions. Social and demographic variables included maternal and household details. The PAediatric Risk Assessment mobile application (*21*) calculated the post-discharge mortality risk score for Phase-2 participants.

Following discharge, field officers contacted caregivers by phone at 2 and 4 months, and in-person at 6 months, to determine vital status, any occurrence of post-discharge health-seeking, and any readmission details.

### Intervention

Phase-2 evaluated the risk-differentiated care package, *Smart Discharges* (**Figure 1**). All children enrolled during this phase were enrolled in the same manner as during Phase-1 by the same research team. Routine clinical care continued as during Phase-1.

**Figure 1.**
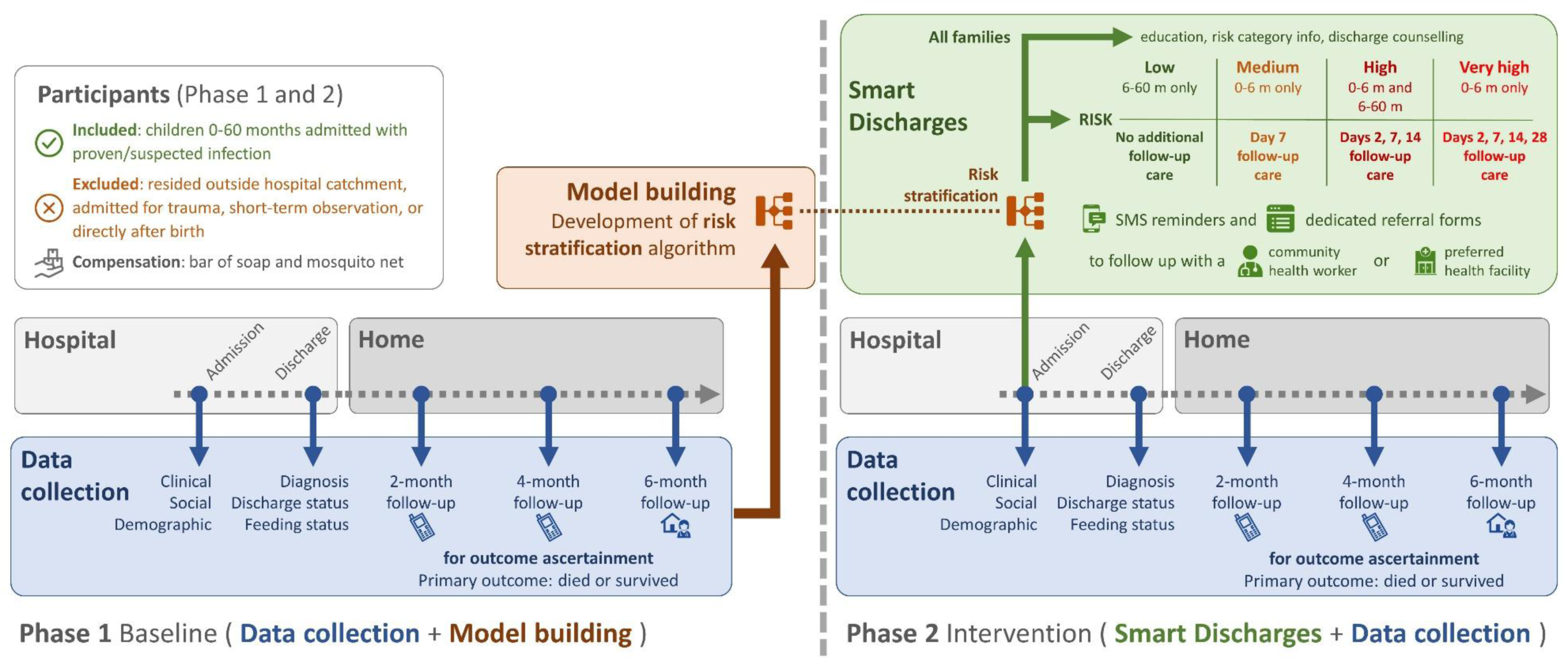
Study overview

At enrollment, children were assessed for risk of post-discharge mortality using predictive algorithms developed from the Phase-1 cohort. The children enrolled in Phase-2 were divided into different risk strata based on age and thresholds for their individual level of risk: low (6-60-months only); medium (0-6-months only); high (0-6-months and 6-60-months); or very high (0-6-months only). These consensus-based divisions balanced the need to have separate models for the two age strata, which have different baseline risks (e.g., no low-risk category for 0-6-months), with the aim of achieving internal consistency for the intensity of the intervention.

At discharge, caregivers of Phase-2 children (regardless of risk category) were told the relative degree of their child’s vulnerability (i.e., the risk category derived from their admission assessments), but not an exact mortality risk score. Caregivers were also given additional counselling and education, including instructional videos, on post-discharge care topics: post-discharge vulnerability, health seeking practices, and general care practices (**Supplementary-S1**). At discharge, caregivers of Phase-2 children were given education materials to take home (**Supplementary-S1**), including a bag with a caregiver guide describing 22 key family care practices, an exercise book where healthcare providers would write clinical notes, and a referral note.

After discharge, families in Phase-2 received a care approach based on their risk category. Children 6-60-months categorized as low-risk received no additional follow-up care. Children 0-6-months categorized as medium-risk received a referral for a single follow-up visit after 7 days. Children of any age with high or very high predicted risk were referred for follow-up care at days 2, 7, and 14 after discharge. In addition, children 0-6-months with very high predicted risk received one additional follow-up referral at 28 days. The proportion of children in Phase-1 who died post-discharge for each risk category is shown in **Supplementary-S3.3**.

Referral forms were made to the community health worker or a facility of choice (emphasis was placed on the family’s choice of a trusted provider or facility and proximity to the home), describing pertinent clinical details of the index admission and a recommendation to assess recovery (**Supplementary-S2**). Automated SMS messages were delivered to caregivers to remind them of their referral, with an additional phone call reminder for infants in the very high-risk category for their first three visits. Our post-discharge referrals did not interfere with any scheduled follow-up arranged by the treating clinicians, who were aware of this project and had access to each enrolled child’s risk details. No monetary or transportation-related support was provided to facilitate follow-up visits.

We recorded adherence to the intervention in Phase-2 by tracking the follow-up referrals made, appointments attended, and reasons for non-attendance. We did not specifically track adverse events or unintended effects of the intervention since we were not providing an intervention exposure with a direct adverse event risk.

### Statistical Analysis

#### Sample size

The sample size was initially determined based on the need to develop prediction models for post-discharge mortality using Phase-1 data (*15*). We sought to enroll cohorts of approximately equal size during Phase-2. Assuming a baseline mortality rate of 5% and a critical alpha of 0.5, this provided 80% power to detect a 20% relative reduction in 6-month all-cause post-discharge mortality. The Phase-1 cohort included additional children because COVID-19 delayed transition of the 0-6-months age group into Phase-2.

#### Statistical methods

Descriptive statistics on demographic, clinical, social, and discharge variables were summarized using median (interquartile range [IQR]) for continuous variables and proportions for categorical variables. Variables were compared between the two phases using the Mann-Whitney U test for continuous variables and Fisher’s exact and Chi-square tests for binary and categorical variables, respectively. We calculated standardized mean difference between phases.

Data were analyzed on an intention-to-treat basis; that is, children were compared between periods, Phase-1 vs. Phase-2, not according to whether they adhered to the intervention. Summary data are given for Phase-2 follow-up referrals made, appointments attended, and reasons for non-attendance.

We assessed the impact of the intervention using an adjusted Cox regression model with post-discharge mortality as the outcome, intervention as the main predictor, and the following covariates, which had been identified *a priori*: age, sex, hospital site, month since first discharge by cohort and phase, and the predicted probability of post-discharge mortality derived from the risk algorithm. We further investigated possible effect modification of the intervention by examining the following subgroups: hospital site; patient ages grouped as 24-60 months, 6-24 months, 2-6 months, <2 months; those with the lowest category anthropometry measures, including mid-upper arm circumference <110 for 6-60 months or <115 for 0-6-months, weight-for-age Z-score <-3, length-for-age Z-score <-3, body mass index Z-score <-3, and weight-for length Z-score <-3; days since discharge of ≤14, ≤30, ≤60, or ≤90 for those who died; clinical presentation of malaria, diarrhea, pneumonia, moderate anemia, or severe anemia; and excluding the COVID period.

Adherence to the intervention was analyzed using descriptive statistics, including reasons for non-attendance. To determine if the number of scheduled visits impacted adherence, we used the Chi-square test to compare the proportion of participants who attended all their scheduled visits in each of the risk categories that have follow-up care referrals (medium, high, and very high). To explore the impact of referral adherence on post-discharge mortality we built a Cox-proportional hazards model, adjusted for mortality risk, using Phase-2 data with mortality as the outcome and adherence as the exposure variable.

Statistical analyses were performed using Stata/MP version 15.0 (StataCorp LLC, College Station, TX) and R version 4.1.3 (R Foundation for Statistical Computing, Vienna, Austria).

## Results

A total of 14,047 (Phase-1, n=7,505; Phase-2, n=6,542) participants were enrolled: 7,100 (94.6%) participants survived to hospital discharge in Phase-1; and 6,282 (96.0%) survived and received the Smart Discharges intervention in Phase-2; the adjusted hazard ratio for in-hospital mortality was 0.77 (95%CI 0.66 to 0.90).

The primary outcome was ascertained from 6,955 (98.0%) participants in Phase-1, who completed follow-up data collection over the six-month post-discharge period, and from 6,096 (97.0%) participants in Phase-2 (**Figure 2**). The characteristics of children lost to follow-up were largely similar to those with completed data (**Supplementary-S3.1**).

**Figure 2.**
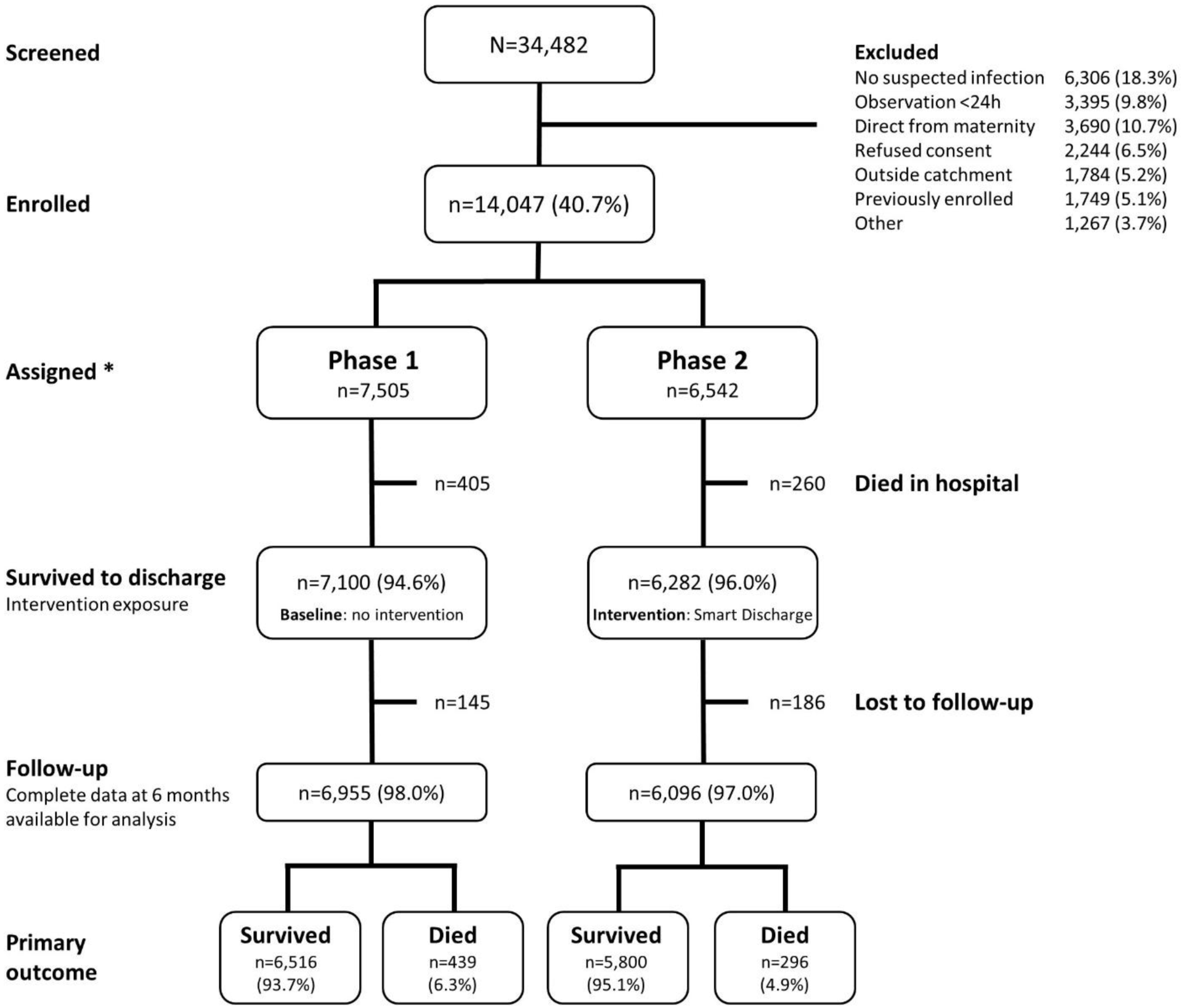
Flowchart showing study enrolment, intervention assignment, and follow-up * Due to the emergence of SARS-CoV-2 in March 2020, the transition of children aged 0-6-months into the intervention Phase-2 was delayed, resulting in 960 additional children being enrolled into the baseline Phase-1.

Overall, 56% of participants in both phases were male; the median (IQR) ages were 0.6 (0.1-1.5) years in Phase-1 and 0.8 (0.2-1.8) years in Phase-2 (**Table 1**) with a standard mean difference of 0.17 years.

**Table 1.** Participant characteristics in the two study phases.

|  | Phase-1<br>(N=6,955) | Phase-2<br>(N=6,096) | P-value | Standardized<br>mean<br>difference |
| --- | --- | --- | --- | --- |
| **a) Demographic variables** |  |  |  |  |
| Male Sex | 3,882 (55.8%) | 3,397 (55.7%) | 0.93 | 0.002 |
| Age (years) | 0.6 (0.1–1.5) | 0.8 (0.2 – 1.8) | <0.001 | -0.17 |
| **Admission Anthropometry** |  |  |  |  |
| Mid Upper Arm Circumference (mm) | 129 (111-142) | 130 (115 – 144) | <0.001 | -0.10 |
| Weight for age Z-scores | -0.94 (-2.1 to 0.01) | -1.1 (-2.1 to -0.1) | 0.02 | 0.03 |
| Length for age Z-scores | -0.7 (-2.0-0.5) | -0.9 (-2.1 – 0.3) | <0.001 | 0.09 |
| BMI Z-scores | -0.9 (-2.2-0.3) | -0.9 (-2.0 – 0.2) | 0.05 | -0.04 |
| Weight for length Z-scores | -0.9 (-2.3–0.3) | -0.9 (-2.0 – 0.2) | 0.32 | -0.02 |
| **b) Admission Clinical Assessment** |  |  |  |  |
| **How long since last admission** |  |  |  |  |
| never | 4,865 (70.0%) | 4,182 (68.6%) | 0.01 | 0.03 |
| <1 month | 739 (10.6%) | 680 (11.2%) |  | 0.02 |
| 1 month – 1 year | 1,076 (15.5%) | 971 (15.9%) |  | 0.01 |
| >1 year | 275 (3.9%) | 253 (4.2%) |  | 0.01 |
| Care previously sought for current illness | 4,529 (65.1%) | 4,050 (66.4%) | 0.11 | 0.03 |
| **Self-reported poor prior health** |  |  |  |  |
| Good health prior to this illness | 6,021 (86.7%) | 5,206 (85.4%) | 0.05 | 0.03 |
| <1 week | 217 (3.1%) | 236 (3.9%) |  | 0.04 |
| 1 week – 1 month | 400 (5.8%) | 387 (6.4%) |  | 0.03 |
| 1 month – 1 year | 317 (4.6%) | 267 (4.4%) |  | 0.009 |
| Referral | 2,118 (30.4%) | 2,115 (34.7%) | <0.001 | 0.09 |
| Prior antibiotic use | 2,995 (43.1%) | 2,567 (42.1%) | <0.001 | 0.02 |
| Prior antimalaria use | 1,459 (21.0%) | 1,365 (22.4%) | <0.001 | 0.03 |
| SpO<sub>2</sub> (%) | 97 (93–99) | 98 (95-99) | <0.001 | -0.29 |
| Heart Rate (bpm) | 146 (132–161) | 146 (133 – 160) | 0.27 | -0.03 |
| Respiratory Rate (bpm) | 49 (39–62%) | 47 (38 – 58) | <0.001 | 0.12 |
| Systolic BP (mmHg) | 92 (81–101) | 92 (83 – 100) | 0.84 | -0.001 |
| Diastolic BP (mmHg) | 51 (42–59) | 53 (46 – 61) | <0.001 | -0.20 |
| Temp (°C) | 37.2 (36.7-38) | 37.2 (36.8-38) | 0.58 | 0.003 |
| Respiratory distress | 1,278 (18.4%) | 945 (15.5%) | <0.001 | 0.08 |
| Capillary refill time ≥3 seconds | 811 (11.7%) | 792 (13.0%) | 0.02 | 0.04 |
| Abnormal Blantyre Coma Scale | 561 (8.1%) | 389 (6.4%) | <0.001 | 0.07 |
| **Symptoms during the course of the illness** |  |  |  |  |
| Cough <14 days | 4,041 (58.1%) | 3,451 (56.6%) | 0.09 | 0.03 |
| Diarrhoea <14 days | 1,987 (28.6%) | 1,460 (24.0%) | <0.001 | 0.11 |
| Diarrhoea >14 days | 161 (2.3%) | 118 (1.9%) | 0.14 | 0.03 |
| Fever <7 days | 5,677 (81.6%) | 5,124 (84.1%) | <0.001 | 0.06 |
| Vomiting everything | 1,136 (16.3%) | 1,195 (19.6%) | <0.001 | 0.09 |
| Abnormally sleepy | 1,264 (18.2%) | 772 (12.7%) | <0.001 | 0.15 |
| Swelling of both feet | 306 (4.4%) | 340 (5.6%) | 0.002 | 0.05 |
| Changes in urine colour | 734 (10.6%) | 875 (14.4%) | <0.001 | 0.12 |
| Making less usual than usual | 482 (6.9%) | 574 (9.4%) | <0.001 | 0.09 |
| Seizure | 926 (13.3%) | 831 (13.6%) | 0.60 | 0.009 |
| **Malaria test positive** | 1,391 (20.0%) | 1,827 (30.0%) | <0.001 | 0.23 |
| **HIV rapid diagnostic test positive** | 206 (3.0%) | 94 (1.5%) | <0.001 | 0.10 |
| **Hemoglobin (g/dL)** | 12 (10-14) | 12 (9.3 – 13.3) | <0.001 | 0.17 |
| **Lactate (mmol/L)** | 2.1 (1.4–3.1) | 2.0 (1.4 – 2.9) | <0.001 | 0.10 |
| **Glucose (mmol/L)** | 5.7 (4.7-6.9) | 5.4 (4.4-6.6) | <0.001 | 0.14 |
| **c) Maternal and social characteristics** |  |  |  |  |
| **Distance to facility (km)** | 14.5 (4.6–31.4) | 14.8 (4.8-34.5) | <0.001 | -0.06 |
| **Maternal age (years)** | 26 (23–30%) | 27 (23-31) | <0.001 | -0.07 |
| **Household size** | 5 (3–6) | 5 (4-6) | <0.001 | -0.07 |
| **Maternal education** |  |  |  |  |
| No school/≤P3 | 699 (10.0%) | 519 (8.5%) | 0.02 | 0.05 |
| P4-P7 | 2,829 (40.7%) | 2,442 (40.1%) |  | 0.01 |
| S1-S6 | 2,437 (35.0%) | 2,198 (36.1%) |  | 0.02 |
| Post-Secondary | 927 (13.3%) | 880 (14.4%) |  | 0.03 |
| **Maternal HIV status** |  |  |  |  |
| Negative | 6,076 (87.4%) | 5,381 (88.3%) | 0.25 | 0.03 |
| Positive | 549 (7.9%) | 438 (7.2%) |  | 0.03 |
| Unknown | 330 (4.8%) | 277 (4.5%) |  | 0.10 |
| **Bed net use** |  |  |  |  |
| Never | 671 (9.6%) | 769 (12.6%) | <0.001 | 0.09 |
| Sometimes | 514 (7.4%) | 564 (9.3%) |  | 0.07 |
| Always | 5,770 (83.0%) | 4,763 (78.1%) |  | 0.12 |
| **Boil/disinfect/filter water** | 5,057 (72.7%) | 4,183 (68.6%) | <0.001 | 0.09 |
| **d) Discharge characteristics** |  |  |  |  |
| **Length of stay (days)** | 4 (3–6) | 4 (3-6) | <0.001 | 0.05 |
| **Discharge status** |  |  |  |  |
| Routine discharge | 5,876 (84.5%) | 5,488 (90.0%) | <0.001 | 0.17 |
| Referred to higher level of care | 236 (3.4%) | 164 (2.7%) |  | 0.04 |
| Unplanned discharge | 843 (12.1%) | 444 (7.3%) |  | 0.16 |
| **Feeding at discharge** |  |  |  |  |
| Feeding well | 5,986 (86.1%) | 5,603 (91.9%) | <0.001 | 0.19 |
| Feeding poorly | 969 (13.9%) | 463 (7.6%) |  | 0.21 |
| **Predicted risk of mortality, %** | 4.3 (2.9-7.1) | 4.1 (2.7 – 6.5) | <0.001 | 0.06 |

**Note:** Data presented as median (interquartile range) for continuous variables and n (%) for binary variables; significance tested with Mann-Whitney U test for continuous variables, Fisher's exact for binary, and chi-square test for categorical variables. Standardized mean difference is the difference in means for continuous variables and difference in proportions for categorical variables. See also **Supplementary S3.2**, which shows some continuous variables divided into relevant severity categories.

Anthropometry measures were largely similar between phases (the standard mean difference for the Z-scores ranged from 0.02 to 0.10), though Phase-1 had a slightly higher proportion of children classified as having severe malnutrition. There was a slightly higher incidence of hypoxemia in Phase-1, and slightly higher incidences of malaria, severe anemia, and hypoglycemia in Phase-2 (**Table 1**, **Supplementary-S3.2**). Applying the prediction model, the mean predicted risk of post-discharge mortality was 6.3% in Phase-1 vs 5.9% in Phase-2; the median risk was 4.3% and 4.1%, respectively.

The rate of post-discharge mortality was 439/6,955 (6.3%) during Phase-1 versus 296/6,096 (4.9%) during Phase-2, with an adjusted hazard ratio of 0.77 (95%CI 0.67 to 0.90) favouring the intervention (**Figure 3**). Readmissions were recorded for 1,313 (18.9%) children in Phase-1 versus 1,024 (16.8%) children in Phase-2.

**Figure 3.**
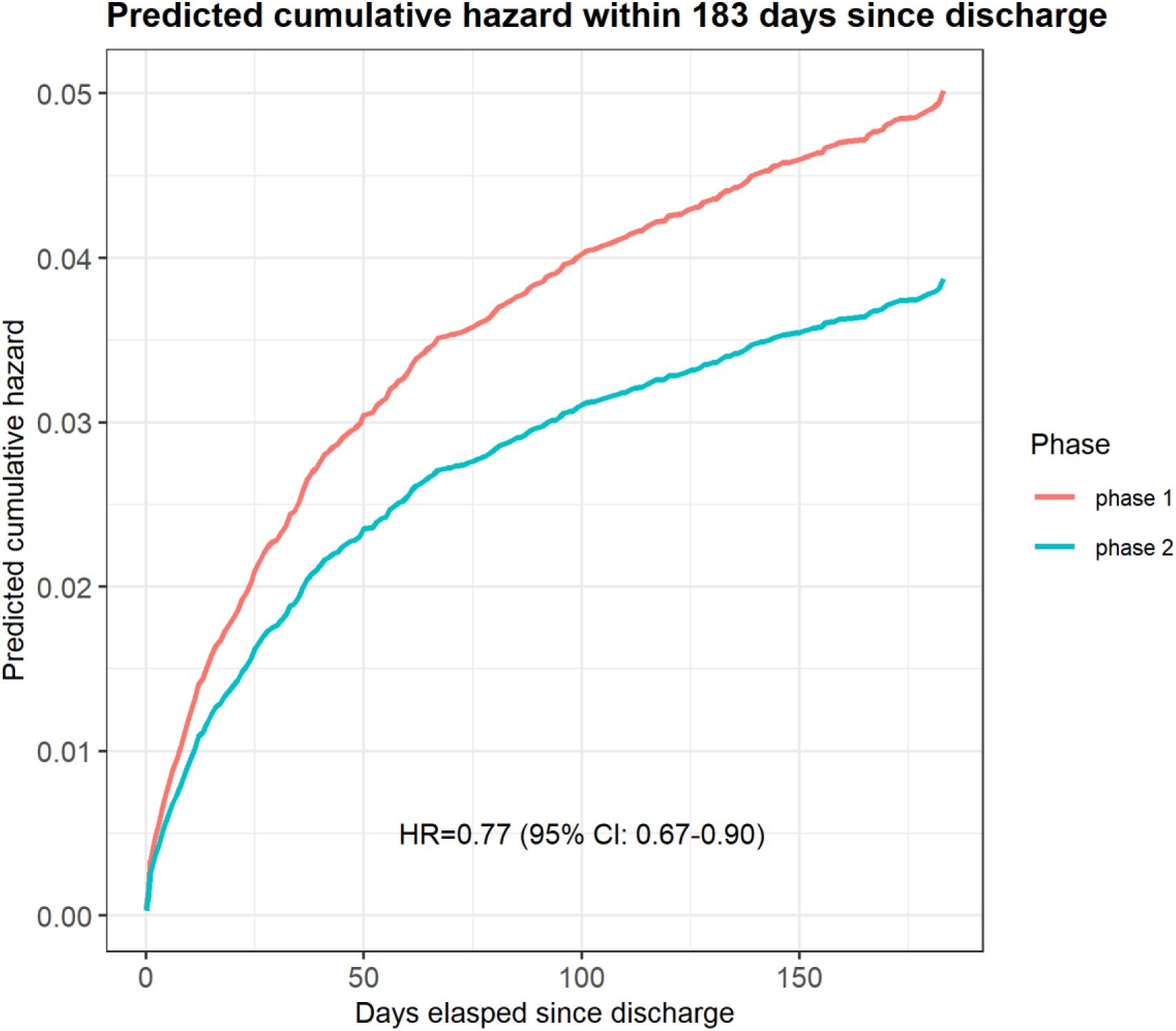
Predicted cumulative hazard from the Cox regression model for mortality in the first 6 months (183 days) following hospital discharge in Phase-1 (baseline) vs Phase-2 (intervention). See also Supplementary-S4.6, which presents raw mortality rate between the phases, with number at risk at each month.

We observed moderate adherence to scheduled follow-up visits, which decreased as the number of assigned visits increased (P<0.001). Of the 6,096 children discharged in Phase-2 with outcomes available: 2,204 were assigned to receive no scheduled follow-up (low risk); 1,205 were assigned a single follow-up (medium-risk) with an adherence rate of 946/1205 (78.5%); 2,466 were assigned three visits (high-risk) with a full adherence rate of 1305/2466 (52.9%); and 220 were assigned four visits (very high-risk, with a full adherence rate of 80/220 (36.4%) (**Table 2**, **Table 3**). Across all risk groups, 931/2,891 (23.9%) participants did not complete any of their scheduled visits. Reasons for non-adherence were most often related to the family considering the referral unimportant as the child was not sick (503/1161, 43.6%) or the visit not being possible for family reasons (286/1161, 24.6%) (**Table 4**). Over 80% of follow-up visits were conducted at heath centres, with fewer than 10% with community health workers. Overall, approximately one-third of follow-up visits resulted in outpatient treatment, referral, or admission.

**Table 2.** Phase-2 follow-up referrals.

| Risk category | Low risk<br>6-60 months only | Medium-risk<br>0-6 months only | High-risk<br>0-6 months and<br>6-60 months | Very high-risk<br>0-6 month only |
| --- | --- | --- | --- | --- |
| **Assigned to risk category**<br>n (% of total N=6,096) | **2,204 (36.2%)** | **1,205 (19.8%)** | **2,467 (40.5%)** | **220 (3.6%)** |
| **Number of referrals assigned** | 0 | 1 | 3 | 4 |
| **Number referrals completed**<br>n (% of column) |  |  |  |  |
| 0 | 2,202 (99.9%) | 253 (21.0%) | 624 (25.3%) | 55 (25.0%) |
| 1 | 1 (0.05%) | 946 (78.5%) | 278 (11.3%) | 15 (6.8%) |
| 2 | 1 (0.05%) | 3 (0.25%) | 260 (10.5%) | 33 (15.0%) |
| 3 | 0 | 3 (0.25%) | 1305 (52.9%) | 37 (16.8%) |
| 4 | - | - | - | 80 (36.4%) |

See also **Supplementary S3.4**, which presents these data split by age cohort (0-6-months vs 6-60-months)

**Table 3.** Phase-2 adherence to intervention.

|  |  | Referral 1 | Referral 2 | Referral 3 | Referral 4 |
| --- | --- | --- | --- | --- | --- |
| **Assigned to follow-up referral** |  | 3,892 | 2,687 | 2,687 | 220 |
| **Attended referral** |  | **2,962 (76.1%)** | **1,722 (64.1%)** | **1,425 (53.0%)** | **80 (36.4%)** |
| **Referral Location**<br>n (% of column) | Community health worker | 239 (8.1%) | 139 (8.1%) | 120 (8.4%) | 8 (10.0%) |
|  | Health centre/clinic | 2,427 (81.9%) | 1,436 (83.4%) | 1,180 (82.8%) | 56 (70.0%) |
|  | Hospital | 259 (8.7%) | 122 (7.1%) | 107 (7.5%) | 16 (20.0%) |
|  | Other | 3 (0.1%) | 2 (0.1%) | 1 (0.1%) | - |
|  | Missing location information | 34 (1.1%) | 23 (1.3%) | 17 (1.2%) | - |
| **Outcome**<br>n (% of column) | No intervention | 1,937 (65.4%) | 1,228 (71.3%) | 1,086 (76.2%) | 72 (90.0%) |
|  | 1. Home-based treatment | 965 (32.6%) | 452 (26.2%) | 293 (20.6%) | 6 (7.5%) |
|  | 2. Admission | 22 (0.7%) | 14 (0.8%) | 17 (1.2%) | - |
|  | 3. Referral | 5 (0.2%) | 8 (0.5%) | 15 (1.1%) | 1 (1.3%) |
|  | **Advice taken n (% of 1-3)** | **983 (99.1%)** | **471 (99.4%)** | **319 (98.2%)** | **6 (85.7%)** |
|  | Missing outcome information | 33 (1.1%) | 20 (1.2%) | 14 (1.0%) | 1 (1.3%) |

Note: N for each column is based on having been assigned to at least that number of referrals; the same children can be included in more than one column. Only medium-, high-, and very high-risk children aged 0-6 months and high-risk children aged 6-60 months are represented in the Table (children assigned to other risk categories were not assigned any referrals).
See also **Supplementary S3.5**, which presents these data split by age cohort (0-6-months vs 6-60-months)

**Table 4.** Reasons for non-adherence, i.e., not attending scheduled appointments.

| **Reasons for non-adherence** | **Age 0-60 month**<br>**(Medium-, High-, or Very High-risk)**<br>**Assigned 1, 3 or 4 referrals**<br>**Did not attend one or more**<br>**(n=1,555)** |
| --- | --- |
| Child not sick and did not consider important | 722 (46.4%) |
| Child away | 30 (1.9%) |
| Child admitted to hospital/health center | 55 (3.5%) |
| Child died | 52 (3.3%) |
| Forgot to go | 156 (10.0%) |
| Visit not possible – health system factor | 121 (7.8%) |
| Visit not possible – family factor | 368 (23.7%) |
| Reason unavailable | 164 (10.5%) |

Note: more than one non-adherence reason could be selected by each family.

In the sensitivity analysis examining hospital site, only one site (the largest in terms of enrollment) demonstrated a significant improvement in hazard ratio for post-discharge mortality between Phase-1 and Phase-2, though the confidence intervals of all sites contained the pooled point estimate, and all but one had a hazard ratio <1.0 (**Figure 4**). Subgroups by anthropometry demonstrated an effect driven by weight-for-age Z-score below −3 and length-for-age Z-score below −3, while the intervention effect by disease group was strongest among children with pneumonia and severe anemia (**Figure 5**). The impact of the intervention on deaths occurring during different time horizons from hospital discharge (14d, 30d, 60d, 90d), had similar, but reduced effect with longer time horizons (**Figure 5**). Further sensitivity analyses of the impact of the intervention on post-discharge mortality using different sets of covariates showed similar results (**Supplementary-S4.2**).

**Figure 4.**
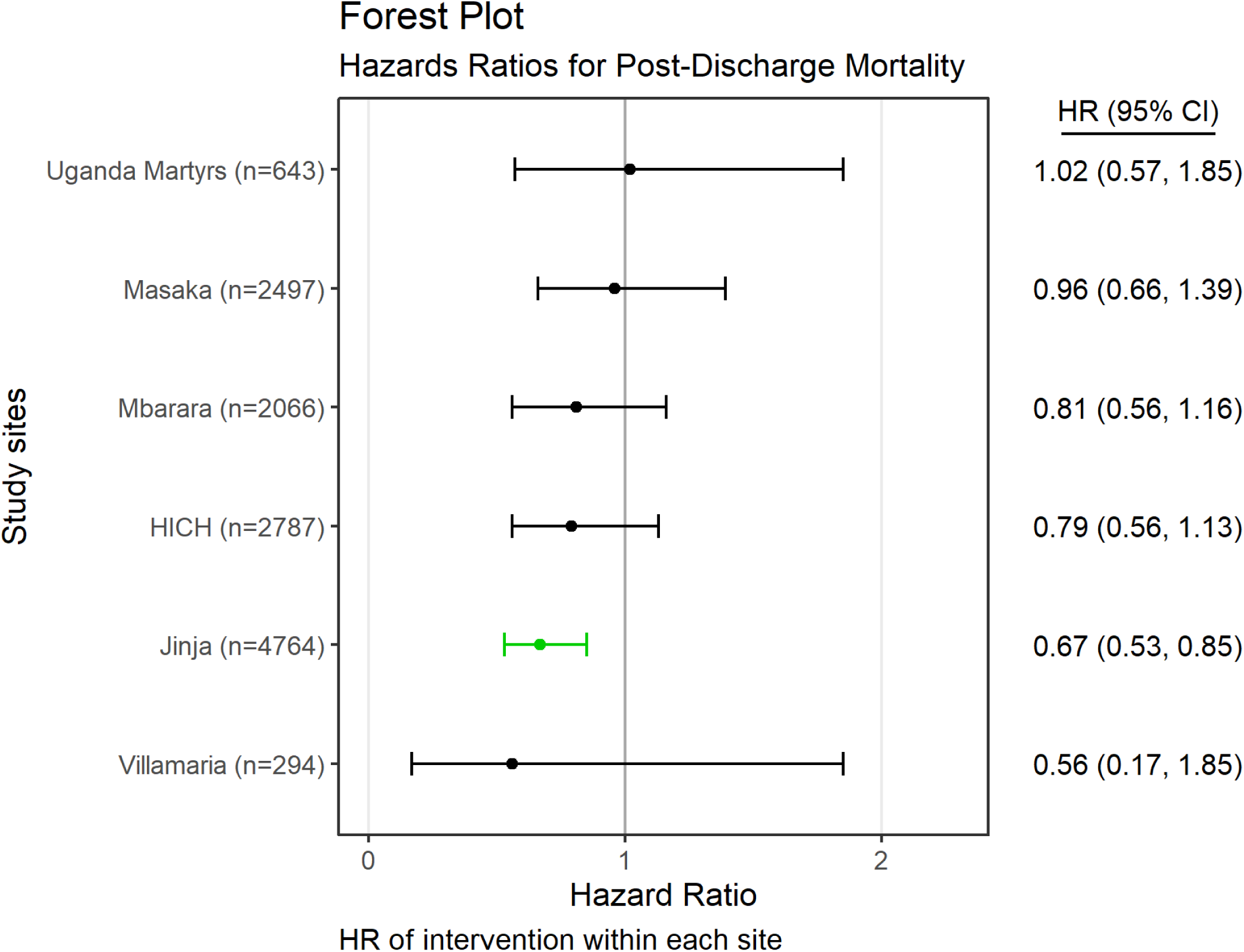
Forest plot showing adjusted hazard ratios for post-discharge mortality at the six different study sites. Adjusted hazard ratios, which are significantly <1, are shown in green.

**Figure 5.**
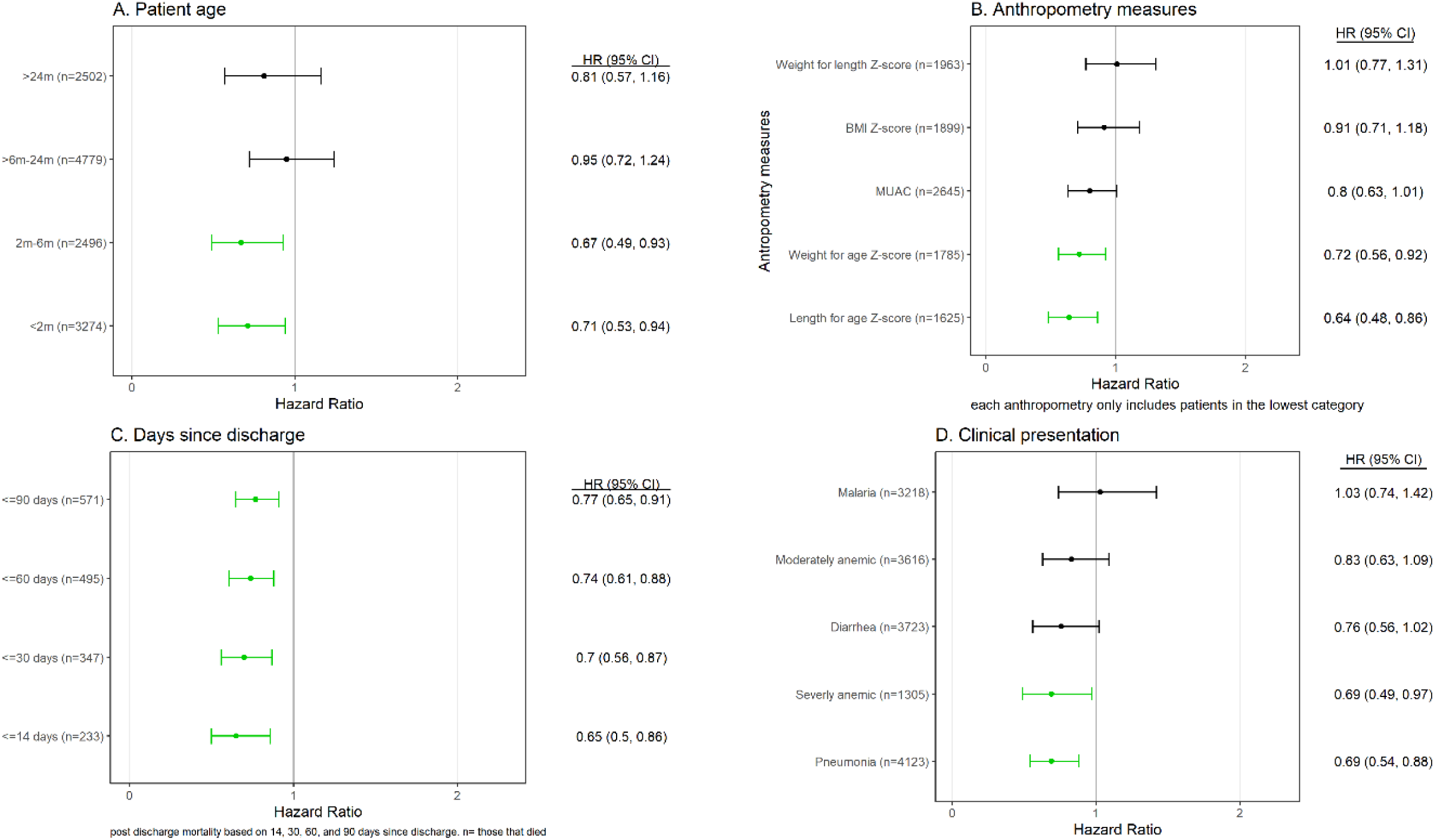
Forest plots showing adjusted hazard ratios for post-discharge mortality based on (A) patient age groups, (B) those with the lowest categories for anthropometry measures, (C) days since discharge for those who died, and (D) clinical presentation. Adjusted hazards ratios, which are significantly <1, are shown in green.

We saw no difference in our pooled estimate in a sensitivity analysis that excluded the 2,516 enrollments between 01-Mar-2020 and 28-Feb-2021, covering the COVID-19 period with the most restrictive transportation limits and the most significant COVID-19 disease peaks (**Supplementary-S4.3**). We observed no association between the predicted risk of mortality and phase (**Supplementary-S4.4**); however, non-adherence to follow-up was associated with a higher risk of post-discharge mortality (**Supplementary-S4.5**).

## Discussion

This study investigated a pragmatic and scalable approach to post-discharge care, demonstrating that a risk-differentiated approach to improving care after discharge in children under five years of age was associated with improved survival. Post-discharge mortality is common among children with acute infectious illness in low-income countries. Deaths after discharge must be addressed if we are to continue to make survival gains in children under five years. These deaths occur in poorly resilient health systems, which are unable to provide post discharge follow-up for all children. In these settings, optimal use of scarce resources dictates that attention be paid to the most vulnerable. Therefore, a precision health approach that focuses on risk assessment, caregiver education and counselling, and increased engagement within the health system during the vulnerable post-discharge period can contribute to achieving child mortality targets of the third Sustainable Development Goal.

Increasingly, digital decision support and risk prediction are being incorporated into clinical care in low resource settings (*22*). Clinical decision support tools are providing transformative opportunities to improve healthcare, often developed using electronic medical record data alongside increasingly sophisticated methods in prediction analysis (*23*). However, the high disease burden and scant data availability has hampered opportunities to improve clinical decision-making in low-income countries (*24*). Yet, these settings represent important opportunities where risk-differentiated approaches for improved decision-making can vastly improve care efficiency and outcomes (*23*). Where the disease burden is high and health worker density is low, approaches to address existing gaps in care, including improving transitions from hospital to home, can more easily be adopted and scaled within a risk-differentiated care framework.

Despite the high burden of pediatric post-discharge mortality, current recommendations by international or national bodies, or by professional associations, do not provide guidance on discharge or post-discharge care within the context of sepsis or acute illness (*8*). This study confirms findings from an earlier feasibility study that identified benefits from increased attention on the discharge process and improved engagement with the health system following hospitalization and discharge among vulnerable children (*13*). A formal assessment of risk by health workers alongside the provision of simple health messages to caregivers related to vulnerability, health-seeking, and health practices provides a starting point to inform improvements in discharge policy and practice. Follow-up visits during the post-discharge period provides further critical support to vulnerable families, as evidenced by about a third of our Phase-2 participants requiring some form of outpatient intervention. This study observed that most post-discharge deaths occurred several weeks after discharge suggesting that engagement beyond the first few weeks may provide additional benefit. Deaths occurring in first 1-2 days following discharge may additionally suggest that data-driven discharge criteria may also support approaches to improve outcomes for children who die shortly after being discharged, although this has not yet been evaluated in these settings. We observed fewer unplanned discharges during implementation, suggesting that improved discharge planning may have also been an important component of the intervention, in addition to the impact of engagement and follow-up following discharge.

A cost-effectiveness analysis is required to better understand the policy-level implications of this intervention, if scaled. However, no financial incentive was provided to families to complete follow-up, and emphasis was placed on local follow-up, either at nearby facilities or with community health workers. Using this approach, adherence was good, with nearly 60% of participants completing all scheduled visits. Furthermore, the use of simple paper-based and digital educational materials, including video-based counselling, is critical to reduce the time required by health workers to provide the patient counselling and education needed for this type of intervention.

### Limitations

This study is subject to several limitations. Most importantly, this study was quasi-experimental and did not randomize either individuals or clusters, such as hospitals. Although the children in the two phases showed similar characteristics, potentially clinically significant differences included higher proportions of children with hypoxemia and severe malnutrition in Phase-1 and higher proportions of children with malaria, severe anemia, and hypoglycemia in Phase-2, which may have impacted survival rates. Consequently, we cannot be confident that temporal confounding did not play a role in the differential results between phases. Several multivariable analyses were conducted, all of which demonstrated similar pooled effects, lending confidence to our interpretation that our estimated interventional effect approximated the true effect.

Secondly, the interventional approach encompassed several concurrent interventions. It is not possible to deduce the components that contributed most to the observed impact. Adherence to follow-up visits was good, but additional strategies to provide a more robust and effective linkage to follow-up care in the community may further improve the intervention’s effectiveness.

Thirdly, the study’s inclusion criteria restricted participants to those with a proven or suspected infectious illness, which does not fully reflect a census sample of all admissions but is most likely to be the target of a potential scaling intervention.

Finally, the contribution of risk prediction itself to the impact of the intervention could not be determined. The knowledge of an objective risk classification could have profound effects on the behavior of clinicians and caregivers, beyond those due to the other aspects of the intervention. With the relative infrequency of risk-differentiated care, it is difficult to know how this approach would work in the absence of formal risk calculations. This has implications for regions where the validity of post-discharge risk prediction is questionable and suggests a need to further develop and validate risk prediction models more widely.

### Future work

Further evaluation of the Smart Discharges program may be required before any regional or national scale-up initiative. A randomized controlled trial would offer the most robust evidence to maximize confidence in the program’s effectiveness, but may not be the most pragmatic approach in terms of time and cost. Such a trial may be based on a cluster randomized stepped-wedge design, with the intervention applied at the site-level, which should overcome concerns about the potential confounding effect of secular variation over time.

Even without robust randomized trial data, our results offer evidence for the importance of improving post-discharge care and sufficient motivation to investigate the integration of such a program into practice. We also need to investigate post-discharge initiatives directed towards community health engagement. Our study was facility-focused: although the referral note had details of the Smart Discharges intervention, the community health workers who provided follow-up care were not given specific training in post-discharge risk-differentiated care.

## Conclusion

Death among children after hospital discharge remains as high as in-hospital deaths in low-income settings. We demonstrated that a simple approach of risk assessment paired with education and engagement following discharge may be an appropriate strategy to further improve child survival. Policies and guidance in these settings must begin to reflect care practices to address post-discharge mortality.

## Data Availability

Study materials including the study protocol, consent form, data collection tools, de-identified participant data, data dictionary, and the analysis code are available on request to the corresponding author (Matthew O. Wiens) or through the published protocol (https://doi.org/10.5683/SP3/QRUMNQ) and dataset (https://doi.org/10.5683/SP3/REPMSY). Owing to the sensitive nature of clinical data, access to the de-identified data is granted on a case-by-case basis and will require the signing of a data sharing agreement.

## Acknowledgements

The study was supported by funding from Grand Challenges Canada (#TTS-1809-1939), Thrasher Research Fund (#13878), BC Children’s Hospital Foundation, and Mining4Life; these funders had no role in the design or conduct of the study.

We would like to acknowledge all past and present members of the Smart Discharges Research Program for their efforts in data collection, administration, logistics support, and all study activities, including but not limited to: Tumwebaze Godfrey, Agaba Collins, Tumukunde Goreth, Naturinda Mackline, Assimwe Abibu, Nakafero Joan, Kiiza Israel, Kitenda Julius, Kamba Ayub, Kuguminkiriza Brenda, Kabajasi Olive, Kembabazi Brenda, Happy Annet, Tusingwire Fredson, Nuwasasira Agaston, Ankatse Christine, Naturinda Rabecca, Nabawanuka Abbey Onyachi, Kamazima Justine, Kairangwa Racheal, Ounyesiga Thomas, Mwoya Yuma, Twebaze Florence, Bulage Mary, Tugumenawe Darius, Tuhame Dyonisius, Twesigye Leonidas, Kamusiime Olivia, Ainembabazi Harriet, Abaho Samuel, Nakabiri Zaituni, Naigaga Shaminah, Kisame Zorah, Babirye Clare, Kayegi Maliza, Opuko Wilson, Mwaka Savio, Baryahirwa Hassan, Mutungi Alexander, Charlene Kanyali, Catherine Kiggundu, Alexia Krepiakevich, Brooklyn Nemetchek, Maryum Chaudhry, Peter Lewis, Rishika Bose, Sahar Zandi Nia, and Tamara Dudley. Without their effort and support, this study would not have been possible.

