## Supplementary for "Smart Discharges improves post-discharge mortality among children with suspected sepsis in Uganda: A prospective before-after study"

### S1 Discharge educational/counselling materials

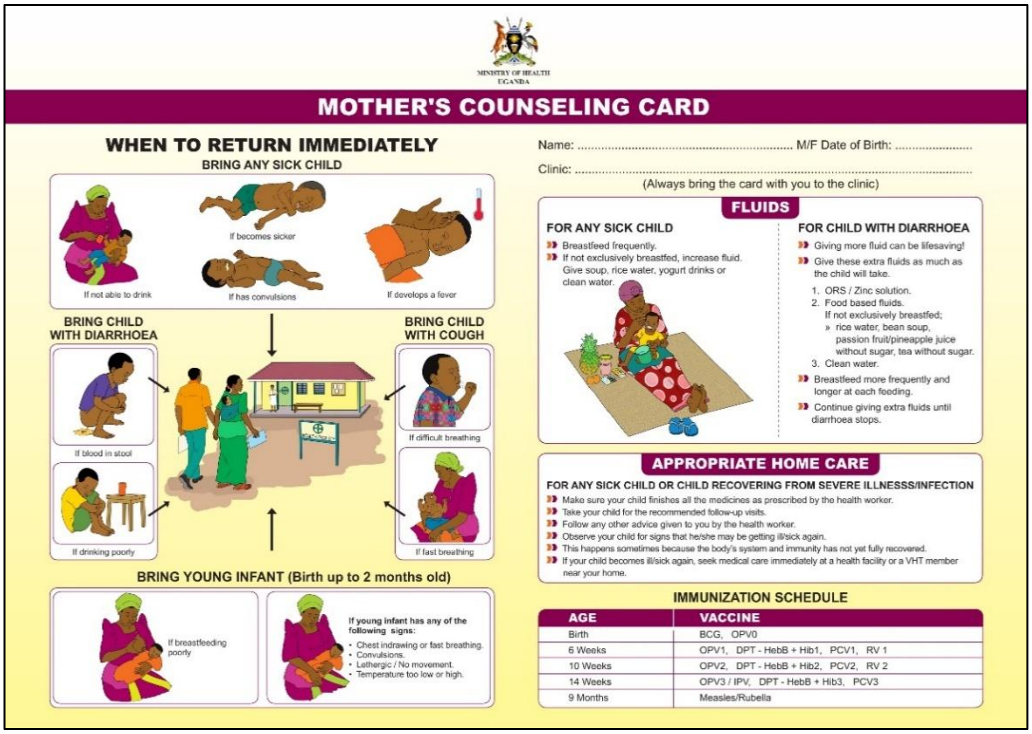

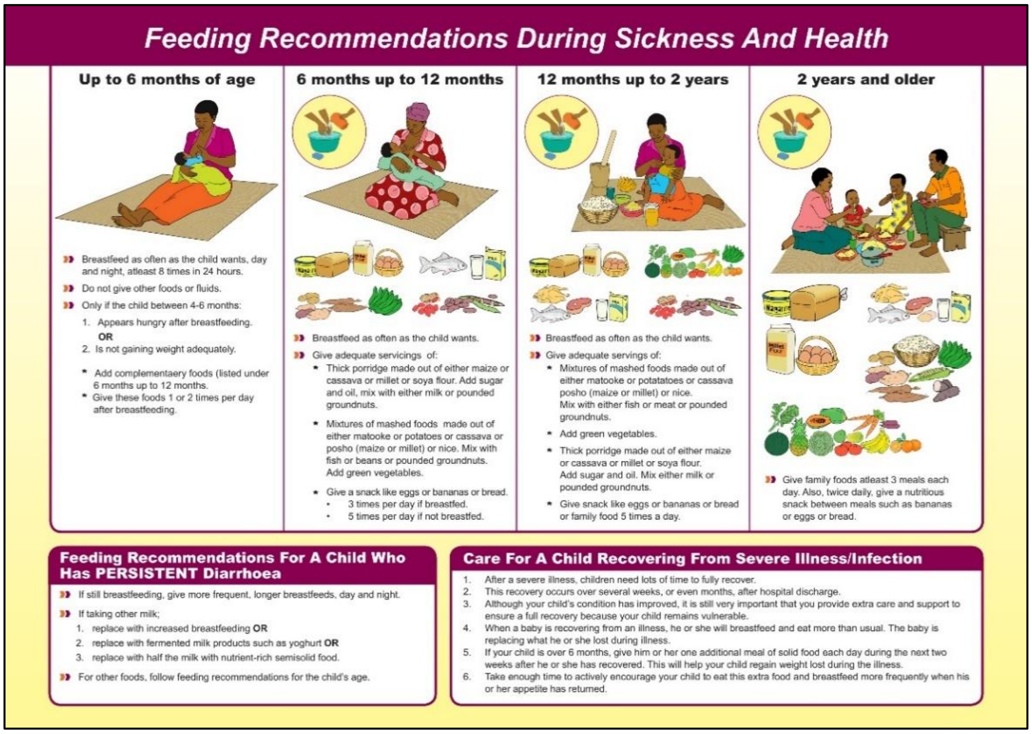

**Figure S1.1** Mother's Counselling Card (2 pages)

Uganda Ministry of Health; Komugisha, Clare; Kenya-Mugisha, Nathan; Wiens, Matthew, 2022, "Counselling Guides for Post-Discharge Care ~ Smart Discharges", https://doi.org/10.5683/SP3/HZ95WN, Borealis, V1

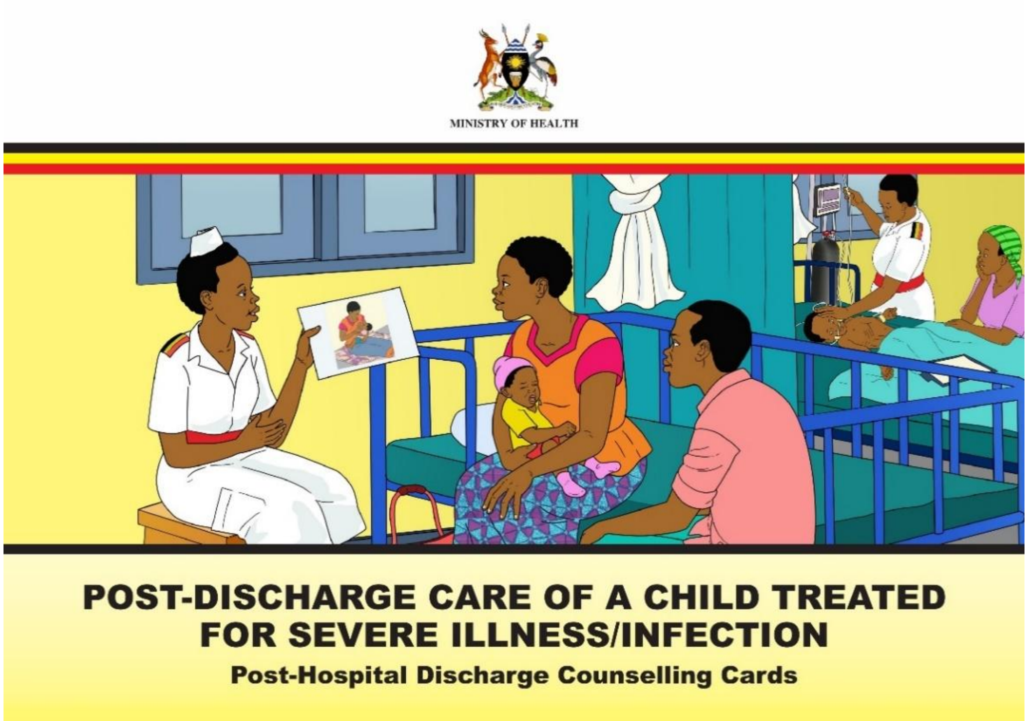

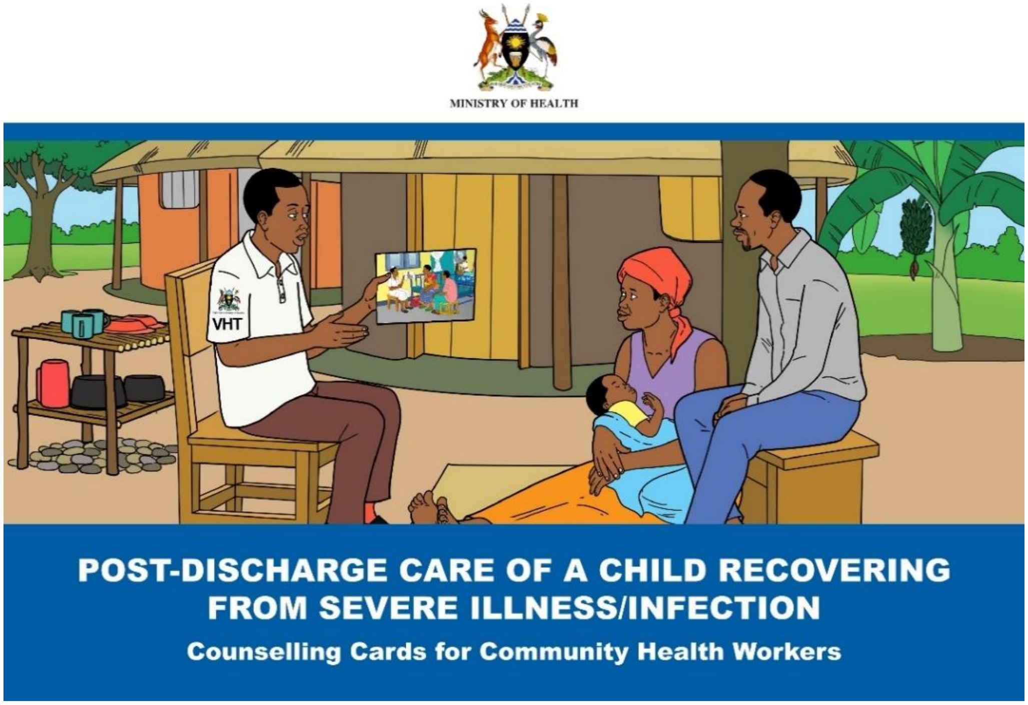

**Figure S1.2** Post-Hospital Discharge Counselling Cards (top, 20 pages) and Counselling Cards for Community Health Workers (bottom, 18 pages)

Uganda Ministry of Health; Komugisha, Clare; Kenya-Mugisha, Nathan; Wiens, Matthew, 2022, "Counselling Guides for Post-Discharge Care ~ Smart Discharges", https://doi.org/10.5683/SP3/HZ95WN, Borealis, V1

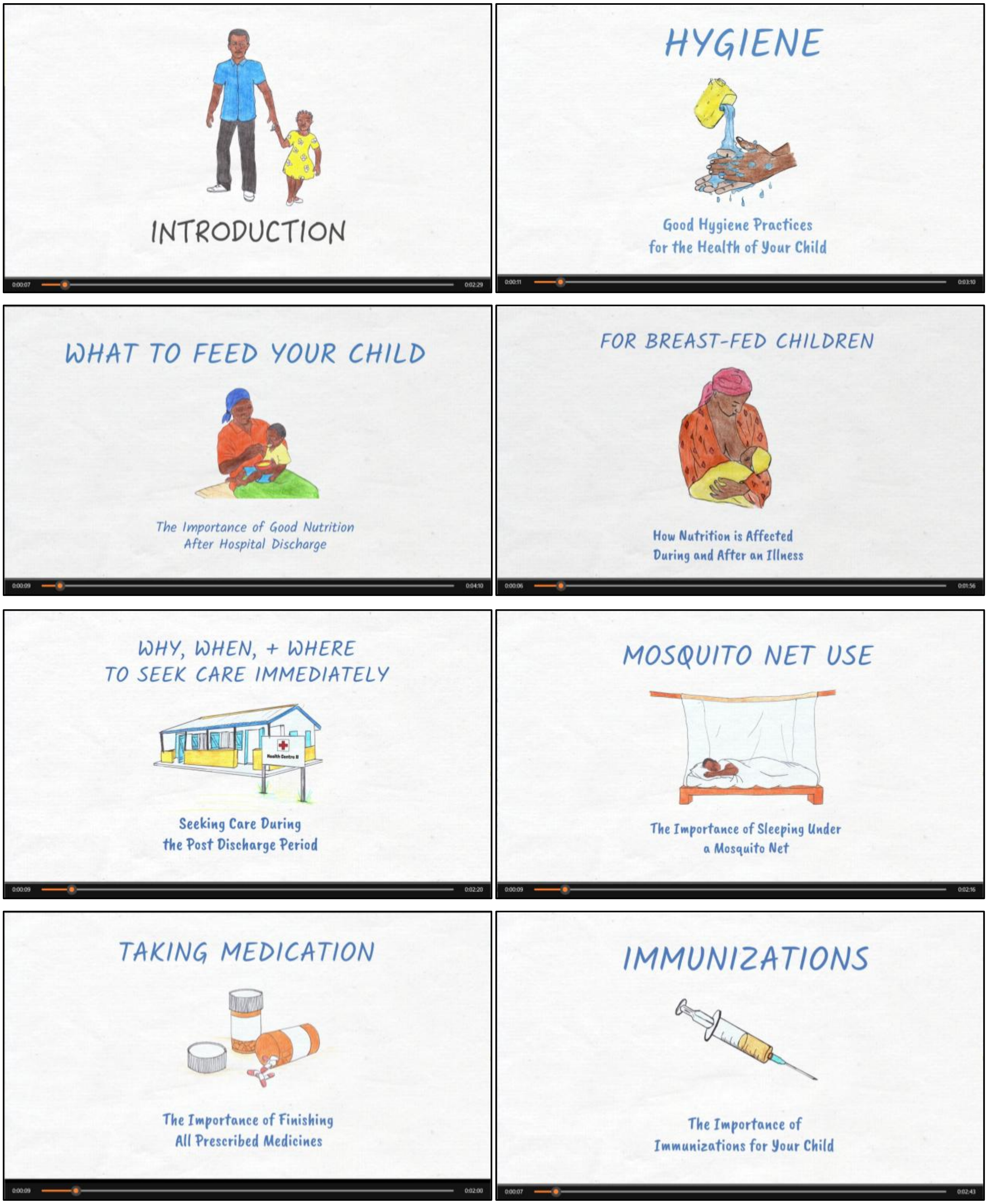

**Figure S1.3** Eight educational videos, each 2-4 minutes long

Komugisha, Clare; Trawin, Jessica; Mwaka, Savio; Kajumba Nsangi, Damalie; Kanyali, Charlene; DeKorne, Micah; Kenya-Mugisha, Nathan; Wiens, Matthew, 2022, "Smart Discharges Uganda Caregiver Counselling Video Series - English Version", https://doi.org/10.5683/SP3/DPQYWC, Borealis, V1

### S2 Data collection materials

**VHT FOLLOW-UP FORM**

**PART I**

**To be filled at the discharging facility and given to the patient / caregiver to give to their VHT for Follow up**

Patient Name: ________________________________ Reference number: ______________________________

Age: ______________________________________ Sex: ☐ Male ☐ Female

Admission Date: ________________________________________ Discharge Date: ________________________

Address: ____________________________________________________________________________________

Discharging Hospital/ Facility___________________________________________________________________

Main Presenting Complaint: _____________________________________________________________________

Discharge Diagnosis: ______________________________ Post Discharge Risk of Mortality: ________________

Referral Health Facility: ____________________________________________ District: _____________________

Name of VHT: _____________________________________________ Telephone Contact: ___________________

Name of the VHT Coordinator: _______________________________ Telephone Contact: ___________________

In- Charge of the Nearest Facility: _____________________________ Telephone Contact: ___________________

Name of Health worker Discharging: ___________________________ Telephone contact: __________________

-----------------------------------------------------------------------------------------------------------

**PART II**

**ATTENTION: Village Health Team Member,**

You are required to follow up this child on the 2nd, 7th and 14th day following discharge from the hospital and offer post discharge care. Examine the child to see if they have any danger signs. Children identified with danger signs should be referred early for further management at the nearest Health Facility.

**Fill out each section of the form for each follow-up appointment.**

---

**Scheduled Visit 1 at day 2:** ____________________ **Actual Visit Date**: ____________________

**Does child have any of the following danger signs?**

☐Vomiting everything ☐Convulsions ☐Chest in-drawing ☐Not able to breast feed or drink

☐Very sleepy/unconscious/difficult to wake ☐Blood in stool ☐Other (specify) __________ ☐None

**Visit Outcome (circle one):**

☐ Counselling on Post discharge care ☐ Referral to the Nearest Facility

VHT Name: ______________________________________________ Signature: _______________________

---

**Scheduled Visit 2 at day 7:** __________________ **Actual Visit Date:** ____________________

**Does child have any of the following danger signs?**

☐Vomiting everything ☐Convulsions ☐Chest in-drawing ☐Not able to breast feed or drink

☐Very sleepy/unconscious/difficult to wake ☐Blood in stool ☐Other (specify) __________ ☐None

**Visit Outcome (circle one):**

☐ Counselling on Post discharge care ☐ Referral to the Nearest Facility

VHT Name: _____________________________________________________ Signature: ____________________

---

**Scheduled Visit 3 at day 14:** ____________________ **Actual Visit Date**: ____________________

**Does child have any of the following danger signs?**

☐Vomiting everything ☐Convulsions ☐Chest in-drawing ☐Not able to breast feed or drink

☐Very sleepy/unconscious/difficult to wake ☐Blood in stool ☐Other (specify) ____________ ☐None

**Visit Outcome (circle one):**

☐ Counselling on Post discharge care ☐ Referral to the Nearest Facility

VHT Name: _____________________________________________________ Signature: ____________________

---

**Figure S2.1** Village Health Team (VHT) follow-up form

Wiens M; Kissoon N; Ansermino JM; Barigye C; Businge S; Kumbakumba E; Larson C; Moschovis P; Singer J; Lavoie P; Kabakyenga J, 2023, "Smart Discharges to improve post-discharge health outcomes in children: A prospective before-after study with staggered implementation", https://doi.org/10.5683/SP3/QRUMNQ, Borealis, V1

### S3 Supplementary tables

**Table S3.1** Participant characteristics of children lost to follow-up in the two study phases

Please compare with Table 1 in the main article.

| | Phase-1 (N=145) | Phase-2 (N=186) |
|---|---|---|
| **a) Demographic variables** | | |
| **Male Sex** | 79 (54.5%) | 85 (45.7%) |
| **Age** (years) | 0.5 (0.1-1.1) | 0.4 (0.1-1.0) |
| **Admission Anthropometry** | | |
| **MUAC** | 125 (115-138) | 122 (105-135) |
| **Weight for age Z-scores** | -0.9 (-2.2 to -0.2) | -1.1 (-2.3 to -0.2) |
| **Length for age Z-scores** | -0.8 (-1.7-0.6) | -0.7 (-1.9-0.6) |
| **BMI Z-scores** | -1.0 (-2.4-0.1) | -1.0 (-2.2 to -0.03) |
| **Weight for length Z-scores** | -1.0 (-2.4-0.3) | -1.2 (-2.3-0.05) |
| **b) Admission Clinical Assessment** | | |
| **How long since last admission** | | |
| never | 99 (68.3%) | 135 (72.6%) |
| <1 month | 17 (11.7%) | 26 (14.0%) |
| 1 month – 1 year | 15 (10.3%) | 23 (12.4%) |
| >1 year | 4 (2.8%) | 2 (1.1%) |
| **Care sought for current illness prior to admission** | 94 (64.8%) | 108 (58.1%) |
| **Self-reported poor prior health** | | |
| Good health prior to this illness | 128 (88.3%) | 162 (87.1%) |
| <1 week | 4 (2.8%) | 9 (4.8%) |
| 1 week – 1 month | 8 (5.5%) | 12 (6.5%) |
| 1 month – 1 year | 4 (2.8) | 3 (1.6%) |
| **Referral** | 43 (29.7%) | 56 (30.1%) |
| **Prior antibiotic use** | 71 (49.0%) | 70 (37.6%) |
| **Prior antimalaria use** | 26 (17.9%) | 23 (12.4%) |
| **$SpO_2$** (%) | 97 (94-99) | 97 (94-99) |
| **Heart Rate** (bpm) | 148 (135-160.5) | 145 (133-158) |
| **Respiratory Rate** (bpm) | 50.5 (39-63) | 54 (42-64) |
| **Systolic BP** (mmHg) | 92 (81-100) | 90 (79-98) |
| **Diastolic BP** (mmHg) | 50 (43-59) | 53 (43-61) |
| **Temp** ($^oC$) | | |
| <36.5 | 20 (13.8%) | 20 (10.8%) |
| 36.5-37.5 | 75 (51.7%) | 103 (55.4%) |
| 37.6-39 | 37 (25.5%) | 51 (27.4%) |
| >39 | 12 (8.3%) | 12 (6.5%) |

| | Phase-1 (N=145) | Phase-2 (N=186) |
|---|---|---|
| **Respiratory distress** | 28 (19.3%) | 35 (18.8%) |
| **Capillary refill time ≥3 seconds** | 9 (6.2%) | 18 (9.7%) |
| **Abnormal Blantyre Coma Scale** | 12 (8.3%) | 15 (8.1%) |
| **Symptoms during the course of the illness** | | |
| Rash | 18 (12.4%) | 3 (1.6%) |
| Cough <14 days | 89 (61.4%) | 105 (56.5%) |
| Cough >14 days | 10 (6.9%) | 10 (5.4%) |
| Diarrhoea <14 days | 50 (34.5%) | 40 (21.5%) |
| Diarrhoea >14 days | 3 (2.1%) | 3 (1.6%) |
| Fever <7 days | 112 (77.2%) | 152 (81.7%) |
| Fever >7 days | 9 (6.2%) | 9 (4.8%) |
| Vomiting everything | 24 (16.6%) | 30 (16.1%) |
| Abnormally sleepy | 36 (24.8%) | 12 (6.5%) |
| Swelling of both feet | 5 (3.5%) | 5 (2.7%) |
| Changes in urine colour | 13 (9.0%) | 16 (8.6%) |
| Making less usual than usual | 9 (6.2%) | 12 (6.5%) |
| Blood in stool | 26 (17.9%) | 1 (0.5%) |
| Seizure | 1 (0.7%) | 32 (17.2%) |
| Coma | 1 (0.7%) | 0 |
| **Malaria test positive** | 31 (21.4%) | 46 (24.7%) |
| **HIV rapid diagnostic test positive** | 8 (5.5%) | 9 (4.8%) |
| **Hemoglobin** (g/dL) | 12 (10.7-14) | 12.7 (10.7-14) |
| **Lactate** (mmol/L) | 2 (1.3-3.1) | 2.1 (1.4-2.8) |
| **Glucose** (mmol/L) | | |
| <2·5 | 4 (2.8%) | 12 (6.5%) |
| 2·5-8·3 | 128 (88.3%) | 157 (84.4%) |
| >8·3 | 13 (9.0%) | 17 (9.1%) |
| **c) Maternal and social characteristics** | | |
| **Time to reach hospital** | | |
| <30 min | 39 (26.9%) | 54 (29.0%) |
| 30 min-1h | 53 (36.7%) | 64 (34.4%) |
| 1-2h | 28 (19.3%) | 40 (21.5%) |
| 2-3h | 18 (12.4%) | 18 (9.7%) |
| > 3h | 6 (4.1%) | 10 (5.4%) |
| **Distance to facility** (km) | 6.3 (4.9-23.1) | 19.3 (10.9-24.4) |
| **Maternal age** (years) | 25 (22-30) | 25 (21-29) |
| **Household size** | 4 (3-5) | 4 (3-6) |
| **Maternal education** | | |
| No school/≤P3 | 21 (14.5%) | 17 (9.1%) |

| | Phase-1 (N=145) | Phase-2 (N=186) |
|---|---|---|
| P4-P7 | 51 (35.2%) | 72 (38.7%) |
| S1-S6 | 56 (38.6%) | 69 (37.1%) |
| Post-Secondary | 16 (11.0%) | 26 (14.0%) |
| **Maternal HIV status** | | |
| Negative | 121 (83.5%) | 160 (86.0%) |
| Positive | 19 (13.1%) | 20 (10.8%) |
| Unknown | 4 (2.8%) | 6 (3.2%) |
| **Bed net use** | | |
| Never | 24 (16.6%) | 30 (16.1%) |
| Sometimes | 10 (6.9%) | 20 (10.8%) |
| Always | 110 (75.9%) | 136 (73.1%) |
| **Boil/disinfect/filter water** | 104 (71.7%) | 149 (80.1%) |
| **d) Discharge characteristics** | | |
| **Length of stay (days)** | 5 (3-6) | 4 (3-7) |
| **Discharge status** | | |
| Routine discharge | 110 (75.9%) | 160 (86.0%) |
| Referred to higher level of care | 4 (2.8%) | 2 (1.1%) |
| Unplanned discharge | 29 (20%) | 24 (12.9%) |
| **Feeding at discharge** | | |
| Feeding well | 50 (34.5%) | 64 (34.4%) |
| Feeding poorly | 95 (65.5%) | 122 (65.6%) |

**Table S3.2** Continuous variables divided into relevant severity categories

| | Phase-1 (N=6,955) | Phase-2 (N=6,096) | P-value | Standardized mean difference |
|---|---|---|---|---|
| **a) Demographic variables** | | | | |
| **Mid Upper Arm Circumference (MUAC)** | | | | |
| <110/<115 | 1,546 (22.2%) | 1,099 (18.0%) | <0.001 | 0.10 |
| 110-120/115-125 | 1,339 (19.3%) | 1,146 (18.8%) | | 0.01 |
| >120/>125 | 4,070 (58.5%) | 3,851 (63.2%) | | 0.10 |
| **Weight for age Z-scores** | | | | |
| <-3 | 954 (13.7%) | 831 (13.6%) | 0.79 | 0.003 |
| -3 to -2 | 922 (13.3%) | 833 (13.7%) | | 0.01 |
| >-2 | 5,079 (73.0%) | 4,432 (72.7%) | | 0.007 |
| **Length for age Z-scores** | | | | |
| <-3 | 838 (12.1%) | 787 (12.9%) | 0.10 | 0.03 |
| -3 to -2 | 861 (12.4%) | 802 (13.2%) | | 0.02 |
| >-2 | 5,256 (75.6%) | 4,507 (73.9%) | | 0.04 |
| **BMI Z-scores** | | | | |
| <-3 | 1,113 (16.0%) | 786 (12.9%) | <0.001 | 0.09 |
| -3 to -2 | 887 (12.8%) | 731 (12.0%) | | 0.02 |
| >-2 | 4,955 (71.2%) | 4,579 (75.1%) | | 0.09 |
| **Weight for length Z-scores** | | | | |
| <-3 | 1,145 (16.5%) | 818 (13.4%) | <0.001 | 0.09 |
| -3 to -2 | 878 (12.6%) | 747 (12.3%) | | 0.01 |
| >-2 | 4,932 (70.9%) | 4,531 (74.3%) | | 0.08 |
| **b) Admission Clinical Assessment** | | | | |
| **$SpO_2$** (%) | | | | |
| $SpO_2$ <90% | 1,061 (15.3%) | 480 (7.9%) | <0.001 | 0.23 |
| $SpO_2$ 90%-95% | 1,656 (23.8%) | 1,195 (19.6%) | | 0.1 |
| $SpO_2$ >95% | 4,238 (60.9%) | 4,421 (72.5%) | | 0.25 |
| **Temp** (°C) | | | | |
| <36.5 | 831 (12.0%) | 587 (9.6%) | <0.001 | 0.07 |
| 36.5-37.5 | 3,406 (49.0%) | 3,206 (52.6%) | | 0.07 |
| 37.6-39 | 2,179 (31.3%) | 1,894 (31.1%) | | 0.006 |
| >39 | 539 (7.8%) | 409 (6.7%) | | 0.04 |
| **Hemoglobin** (g/dL) | | | | |
| Not anaemic (≥11g/dL) | 4,593 (66.1%) | 3,537 (58.0%) | <0.001 | 0.17 |

| | | | | |
|---|---|---|---|---|
| Mild anaemia (7-10 g/dL) | 1,814 (26.1%) | 1,802 (30.0%) | | 0.08 |
| Severe anaemia (<7 g/dL) | 548 (7.9%) | 757 (12.4%) | | 0.15 |
| **Glucose** (mmol/L) | | | | |
| <2·5 | 210 (3.0%) | 364 (6.0%) | <0.001 | 0.14 |
| 2·5-8·3 | 5,983 (86.0%) | 5,182 (85.0%) | | 0.03 |
| >8·3 | 762 (11.0%) | 550 (9.0%) | | 0.06 |
| **e) Post-discharge characteristics** | | | | |
| **Location of death** | | | | |
| In-hospital | 160 (36.5%) | 110 (37.2%) | 0.04 | 0.004 |
| In-transit | 80 (18.2%) | 72 (24.3%) | | 0.14 |
| At home | 193 (44.0%) | 114 (38.5%) | | 0.12 |

**Note**: Data are presented as median (interquartile range) for continuous variables and n (%) for binary variables; statistical significance tested with Mann-Whitney U test for continuous variables, Fisher's exact for binary, and chi-square test for categorical variables.

**Table S3.3** Proportion of children in Phase-1 who died post-discharge for each risk category

| Risk category | 0-6-months | | 6-60-months | |
|---|---|---|---|---|
| | **Died post-discharge, n (% of N)** | **N (% of total)** | **Died post-discharge, n (% of N)** | **N (% of total)** |
| Low | | | 46 (2.1%) | 2,212 (61.3%) |
| Moderate | 47 (3.1%) | 1,521 (45.4%) | | |
| High | 122 (8.7%) | 1,402 (41.9%) | 130 (9.3%) | 1,395 (38.7%) |
| Very high | 94 (22.1%) | 425 (12.7%) | | |
| Total | | 3,348 | | 3,607 |

**Table S3.4** Phase 2 follow-up referrals, age groups separated

| Risk category | **Low risk** 6-60 months only | **Medium-risk** 0-6 months only | **High-risk** 0-6 months and 6-60 months | **Very high-risk** 0-6 month only |
|---|---|---|---|---|
| **Number of referrals assigned** | 0 | 1 | 3 | 4 |
| **0-6-month** **Assigned to risk category** n (% of total **N=2,391**) | | 1205 (50.4%) | 966 (40.4%) | 220 (9.2%) |
| **Number referrals completed** n (% of column) | | | | |
| 0 | | 253 (21%) | 252 (26.1%) | 55 (25.0%) |
| 1 | | 946 (78.5%) | 138 (14.3%) | 15 (6.8%) |
| 2 | | 3 (0.3%) | 87 (9.0%) | 33 (15.0%) |
| 3 | | 3 (0.3%) | 489 (50.6%) | 37 (16.8%) |
| 4 | | - | - | 80 (36.4%) |
| **6-60-month** **Assigned to risk category** n (% of total **N=3,705**) | 2204 (59.5%) | | 1501 (40.5%) | |
| **Number referrals completed** n (% of column) | | | | |
| 0 | 2202 (99.9%) | | 372 (24.8%) | |
| 1 | 1 (0.05%) | | 140 (9.3%) | |
| 2 | 1 (0.05%) | | 173 (11.5%) | |
| 3 | 0 | | 816 (54.4%) | |

**Table S3.5** Phase 2 adherence to intervention, age groups separated

| | | Referral 1 | Referral 2 | Referral 3 | Referral 4 |
|---|---|---|---|---|---|
| **0-6-month** | **Assigned to follow-up referral** | 2,391 | 1,186 | 1,186 | 220 |
| | **Attended** | **1,831 (76.6%)** | **732 (61.7%)** | **609 (51.3%)** | **80 (36.4%)** |
| **Referral Location** n (% of column) | Community health worker | 100 (5.5%) | 34 (4.6%) | 32 (5.3%) | 8 (10.0%) |
| | Health centre/clinic | 1,484 (81.1%) | 600 (82.0%) | 499 (81.9%) | 56 (70.0%) |
| | Hospital | 228 (12.5%) | 90 (12.3%) | 72 (11.8%) | 16 (20.0%) |
| | Other | 1 (0.1%) | 2 (0.3%) | 1 (0.2%) | - |
| | Missing | 18 (1.0%) | 6 (0.8%) | 5 (0.8%) | - |
| **Outcome** n (% of column) | No intervention | 1,394 (76.1%) | 607 (82.9%) | 535 (87.9%) | 72 (90.0%) |
| | 1. Home-based treatment | 403 (22.0%) | 107 (14.6%) | 60 (9.9%) | 6 (7.5%) |
| | 2. Admission | 16 (0.9%) | 7 (1.0%) | 4 (0.7%) | - |
| | 3. Referral | - | 4 (0.6%) | 5 (0.8%) | 1 (1.3%) |
| | ***Advice taken*** *n (% of 1-3)* | *414 (98.8%)* | *117 (99.2%)* | *66 (95.7%)* | *66 (82.5%)* |
| | Missing | 18 (1.0%) | 7 (1.0%) | 5 (0.8%) | 1 (1.3%) |
| **6-60-month** | **Assigned to follow-up referral** | 1,501 | 1,501 | 1,501 | |
| | **Attended** | **1,131 (75.3%)** | **990 (66.0%)** | **816 (54.4%)** | |
| **Referral Location** n (% of column) | Community health worker | 139 (12.3%) | 105 (10.6%) | 88 (10.8%) | |
| | Health centre/clinic | 943 (83.4%) | 836 (84.4%) | 681 (83.5%) | |
| | Hospital | 31 (2.7%) | 32 (3.2%) | 35 (4.3%) | |
| | Other | 2 (0.2%) | 0 | 0 | |
| | Missing | 16 (1.4%) | 17 (1.7%) | 12 (1.5%) | |
| **Outcome** n (% of column) | No intervention | 543 (48.0%) | 621 (62.8%) | 551 (67.5%) | |
| | 1. Home-based treatment | 562 (49.7%) | 345 (34.9%) | 233 (28.6%) | |
| | 2. Admission | 6 (0.5%) | 7 (0.7%) | 13 (1.6%) | |
| | 3. Referral | 5 (0.4%) | 4 (0.4%) | 10 (1.2%) | |
| | ***Advice taken*** *n (% of 1-3)* | 569 (99.3%) | 354 (99.4%) | 253 (98.8%) | |
| | Missing | 15 (1.3%) | 13 (1.31%) | 9 (1.1%) | |

Note: N for each column is based on having been assigned to at least that number of referrals; the same children can be included in more than one column. Only medium-, high-, and very-high risk children aged 0-6 months and high-risk children aged 6-60 months are represented in the Table (children assigned to other risk categories were not assigned any referrals).

### S4 Supplementary figures

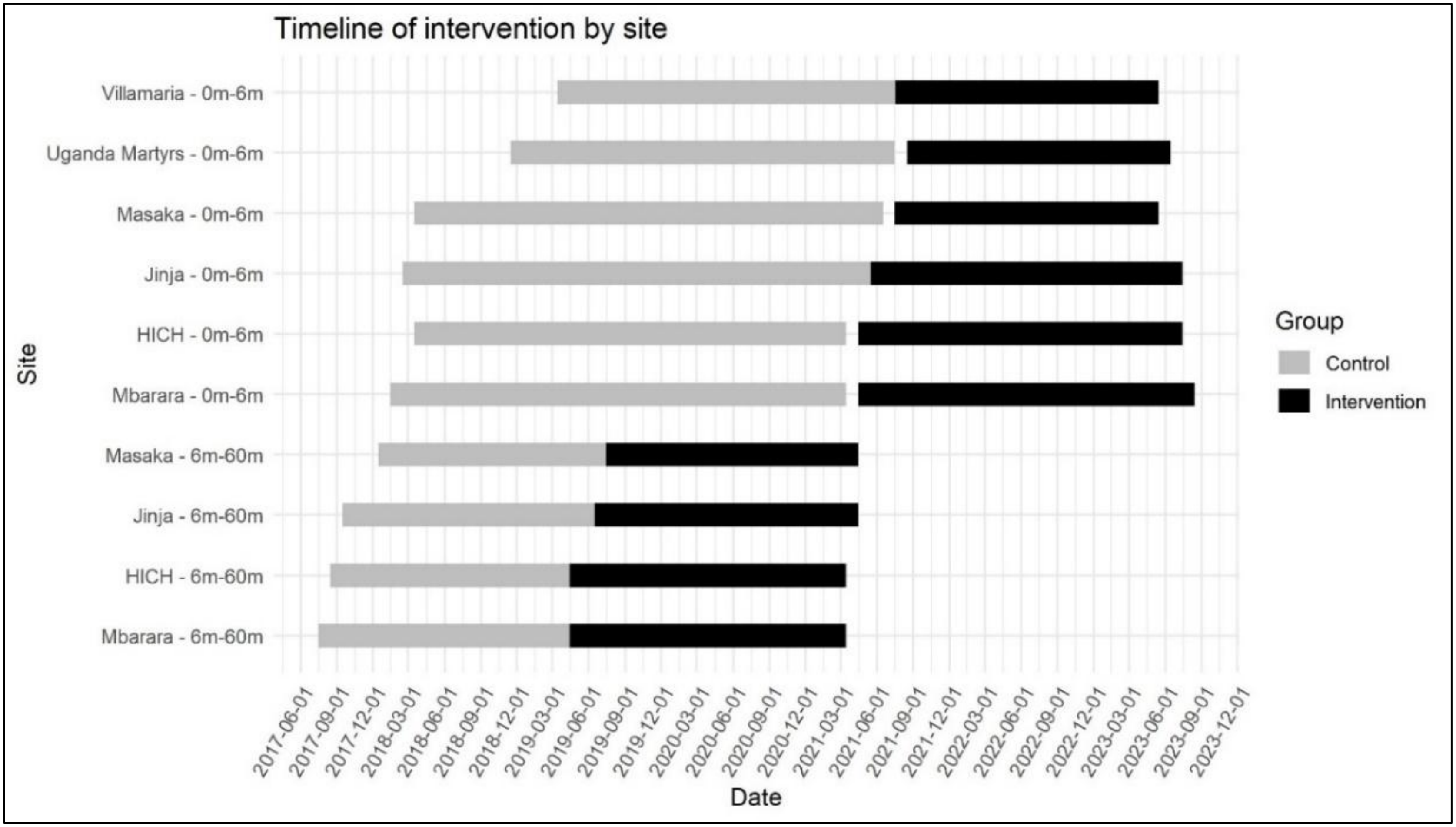

**Figure S4.1 Timeline of baseline (control) and intervention periods by site**

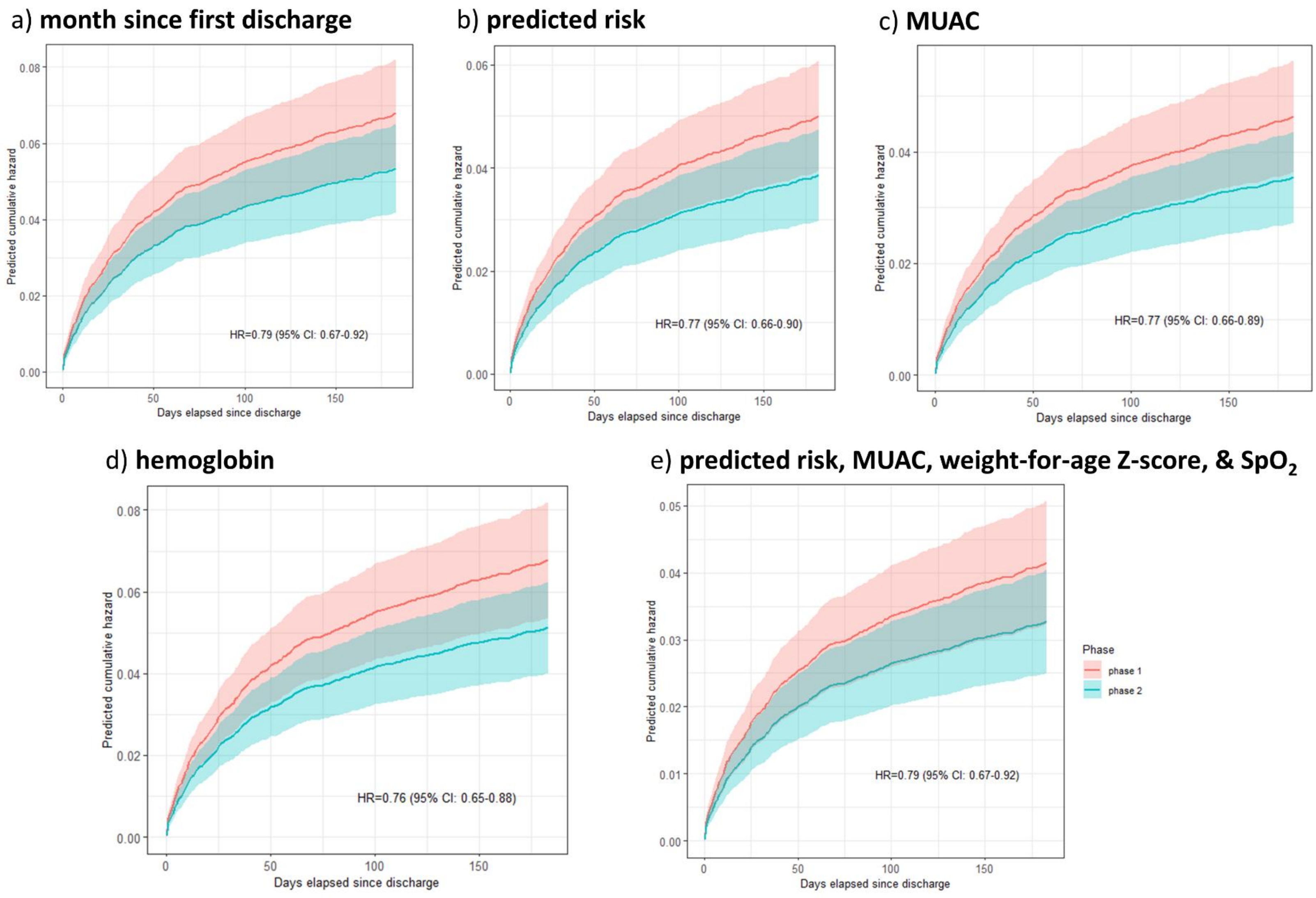

**Figure S4.2** Sensitivity analyses of the impact of the intervention on post-discharge mortality using different sets of covariates

Figure panels [a-e] show predicted cumulative hazard from the Cox regression model for mortality in the first 6 months following hospital discharge in Phase-1 (baseline) vs Phase-2 (intervention); covariates include sex, age, site for all models, along with **other listed covariate(s)**.

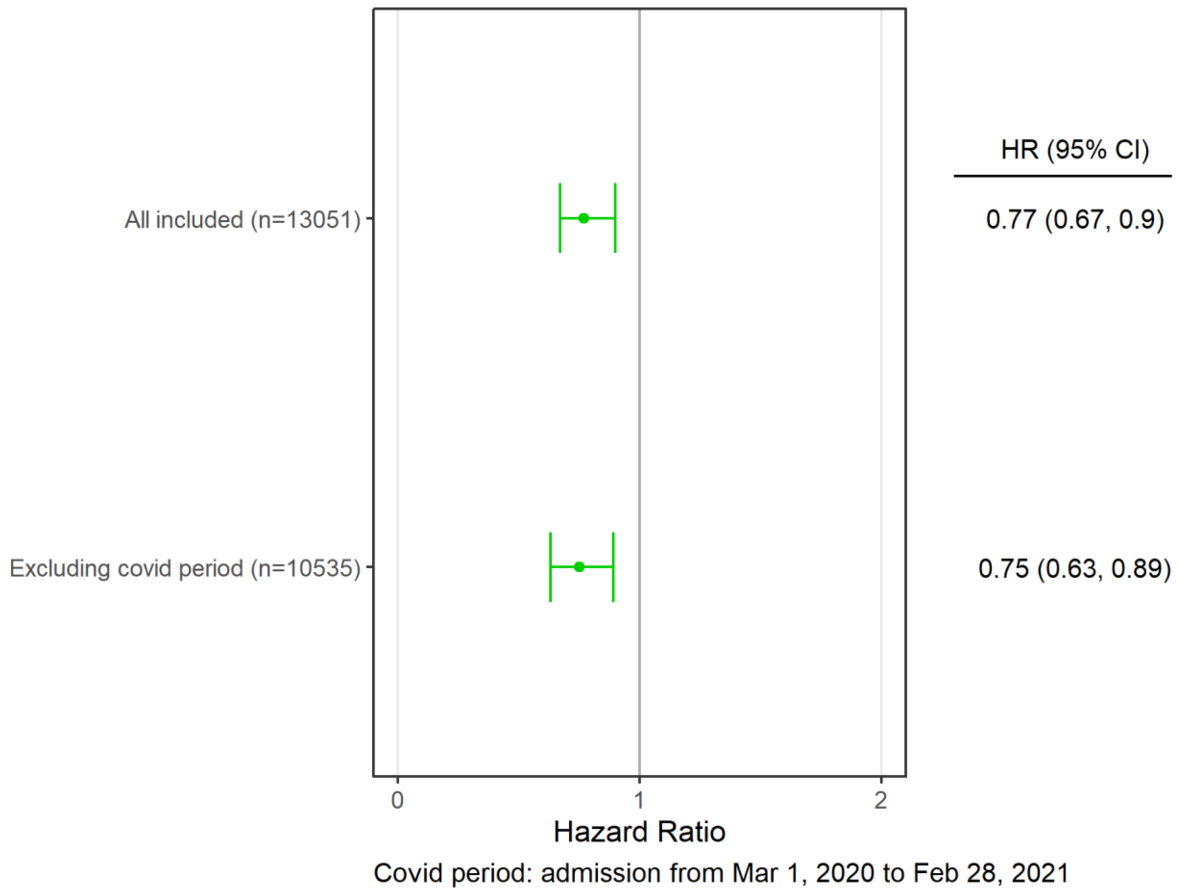

**Figure S4.3** Forest plot showing hazard ratios for post-discharge mortality comparing all participants (COVID period included) vs. the subset of participants excluding the COVID period.

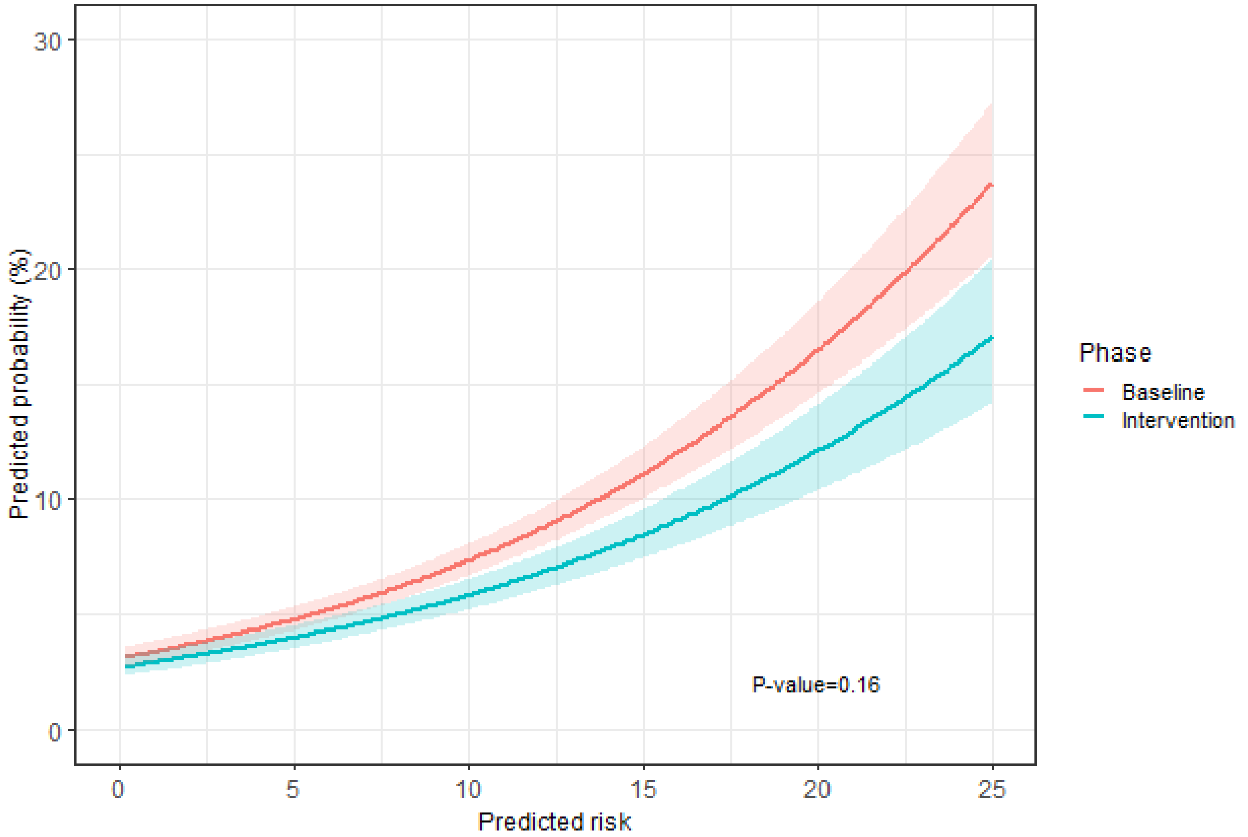

**Figure S4.4** Interaction between predicted risk of mortality and phase.

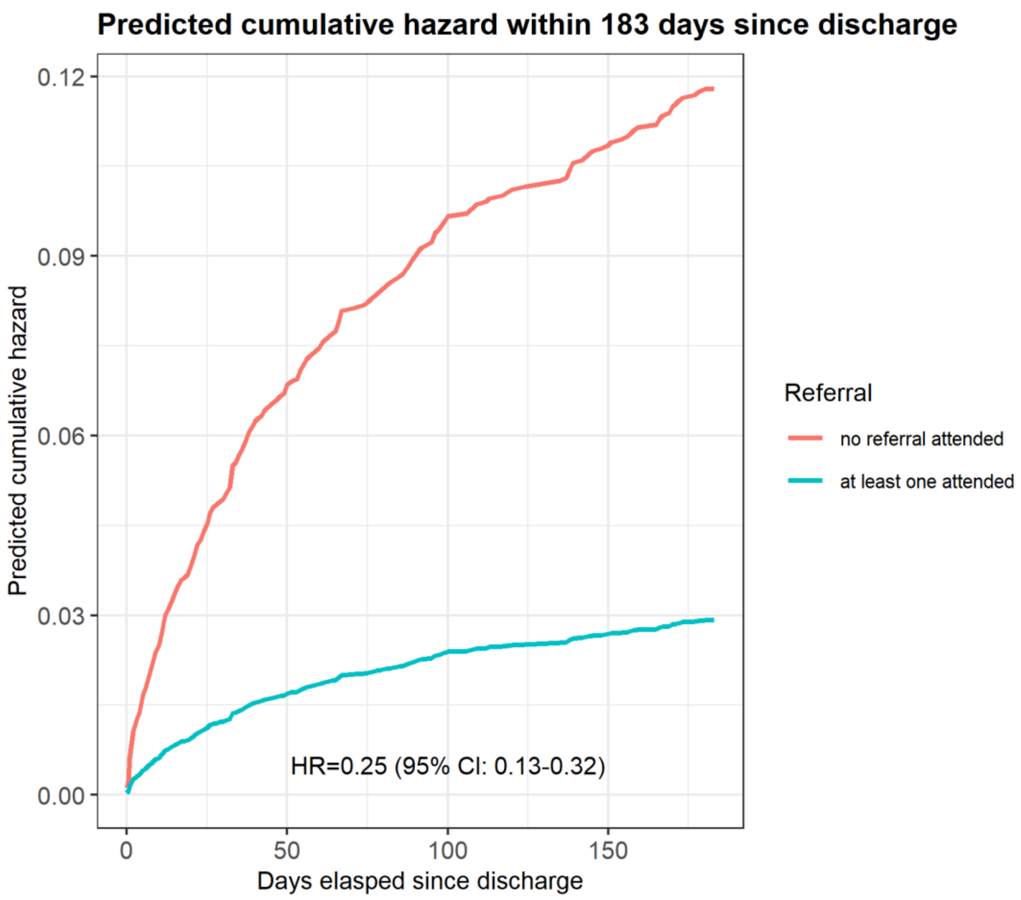

**Figure S4.5** Predicted cumulative hazard for mortality in the first 6 months following hospital discharge.

Stratified by children who attended at least one referral vs. attended no referral. Limited to medium, high, and very high-risk Phase-2 children assigned one or more referrals.

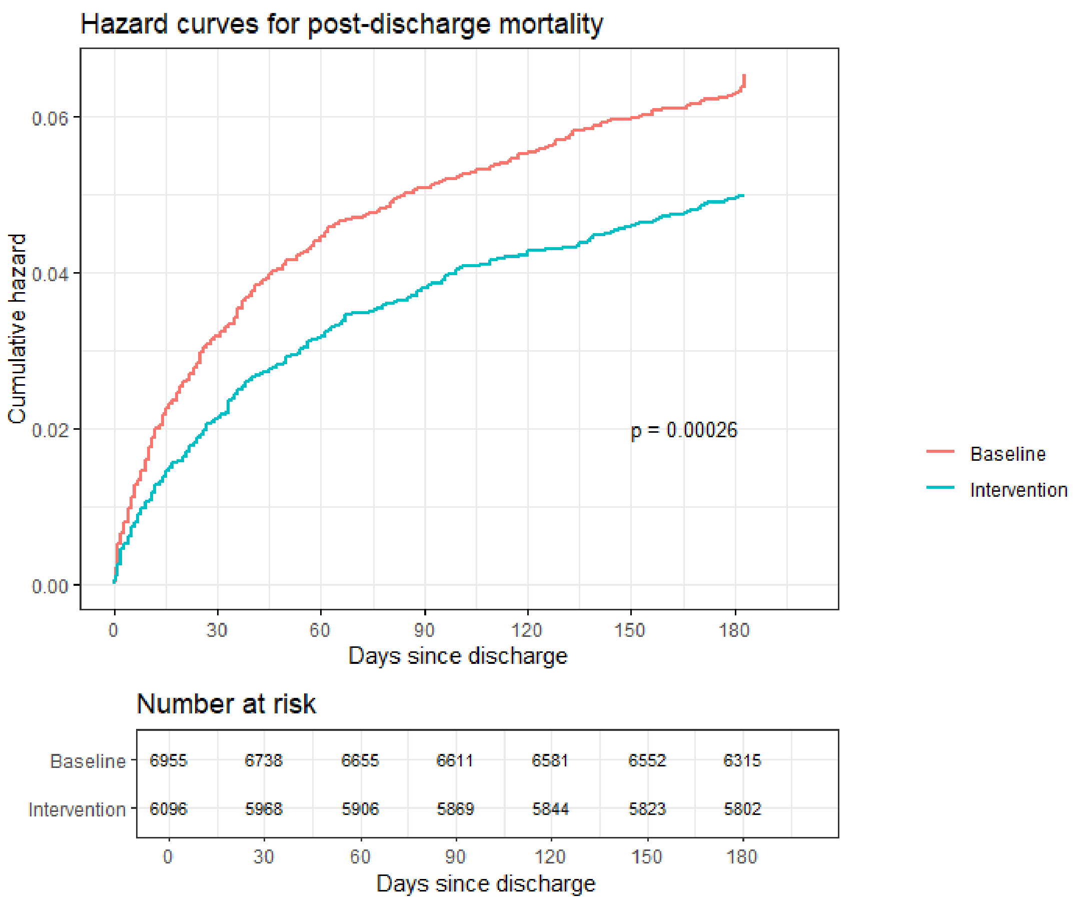

**Figure S4.6** Cumulative hazard for mortality in the first 6 months following hospital discharge.

Stratified by study phase and showing number of children at risk at each month.
